# Impacts of school closures on physical and mental health of children and young people: a systematic review

**DOI:** 10.1101/2021.02.10.21251526

**Authors:** Russell Viner, Simon Russell, Rosella Saulle, Helen Croker, Claire Stansfeld, Jessica Packer, Dasha Nicholls, Anne-Lise Goddings, Chris Bonell, Lee Hudson, Steven Hope, Nina Schwalbe, Anthony Morgan, Silvia Minozzi

**Affiliations:** UCL Great Ormond St. Institute of Child Health, London; Department of Epidemiology, Lazio Regional Health Service, Rome; UCL Institute of Education, London; Division of Brain Sciences, Imperial College London; London School of Hygiene and Tropical Medicine, London; Heilbrunn Department of Population and Family Health, Columbia University, New York; Glasgow Caledonian University, London

## Abstract

**Background:** The well-documented links between education and health mean that school closures during the COVID-19 pandemic are likely to be associated with significant health harms to children and young people (CYP). A systematic review of the evidence is needed to inform policy decisions around school closures and re-openings during the pandemic.

**Methods:** We undertook a high-quality systematic review of observational quantitative studies (published or preprint) of the impacts of school closures (for any reason) on the health, wellbeing and educational outcomes of CYP, excluding impacts of closure on transmission of infection (PROSPERO CRD42020181658). We used a machine learning approach for screening articles, with decisions on inclusion and data extraction performed independently by 2 researchers. Quality was assessed for study type. A narrative synthesis of results was undertaken as data did not allow meta-analysis.

**Results:** 16,817 records were screened, of which 151 were reviewed in full-text and 72 studies were included from 20 countries. 33% were cohort studies using historical control periods; 19% pre-post studies; and 46% cross-sectional studies which assessed change by comparison with population reference data. 63% were high-quality, 25% medium-quality and 13% low-quality. Cause of closure in all studies was the first COVID-19 pandemic wave with the exception of 5 influenza studies and 1 teacher strike.

27 studies concerning mental health identified considerable impacts across emotional, behavioural and restlessness/inattention problems; 18-60% of CYP scored above risk thresholds for distress, particularly anxiety and depressive symptoms. Two studies reported non-significant rises in suicide rates. Self-harm and psychiatric attendances were markedly reduced, indicating a rise in unmet mental health need. Child protection referrals fell 27-39%, with a halving of the expected number of referrals originating in schools.

19 studies concerning health service use showed marked reductions in emergency department (ED) presentations and hospital admissions, with evidence of delayed presentations and potential widening of inequalities in vaccination coverage. Data suggested marked rises in screen-time and social media use and reductions in physical activity however data on sleep and diet were inconclusive. Available data suggested likely higher harms in CYP from more deprived populations.

**Conclusions:** School closures as part of broader social distancing measures are associated with considerable harms to CYP health and wellbeing. Available data are short-term and longer-term harms are likely to be magnified by further school closures. Data are urgently needed on longer-term impacts using strong research designs, particularly amongst vulnerable groups. These findings are important for policy-makers seeking to balance the risks of transmission through school-aged children with the harms of closing schools.

## Background

Nearly every country in the world implemented school closures during 2020 as part of national social distancing efforts to reduce the transmission of SARS-CoV-2 during the COVID-19 pandemic. In March to May 2020, up to 1.5 billion children and young people (CYP) were out of school^1^ in order to reduce social mixing between CYP, disrupt transmission of SARS-CoV-2, and reduce introduction of the virus into households.^2^ Yet almost a year later, the effectiveness of school closures in reducing community transmission continues to unclear,^3^ with high quality studies ranging from no effect^4,5^ through to substantial protective effects.^6,7^

Many countries have closed schools again in early 2021 in response to winter peaks of infection in the northern hemisphere, exacerbated by heightened infectiousness of new virus variants. The benefits of school closures must be considered alongside any harms, both for CYP but also for their families and broader society. The economic harms of school closures through parent work-absenteeism are well documented in the influenza literature.^8,9^ Yet the historical literature provides little guide to the likely health impacts on CYP of widespread and lengthy school closures used to combat COVID-19.^10^

There are strong theoretical reasons to be concerned about the impacts of widespread school closures. Education is one of the strongest determinants of health^11^ and disruption to it influence health and wellbeing in various ways. There is clear evidence that education loss leads to long-term reductions in health and life-expectancy.^12^ Other mechanisms through which school closures may influence CYP health and wellbeing, include isolation of CYP from social support from peers and school staff, loss of school inputs into the provision of health and social care, including child protection notifications and access to mental health support, reduction in physical activity (PA) related to attending school (including but not limited to school sports and exercise) and loss of access to school food programmes for deprived CYP. ^13^

A number of international reports have identified the broad harms done to CYP by social lockdowns and school closures during COVID-19, concluding that responses to the pandemic internationally have worsened CYP outcomes globally, particularly amongst those already disadvantaged.^14^ Yet these efforts have not attempted to examine the impacts of school closures in isolation nor systematically identify all relevant literature. These are needed to inform policy decisions balancing the benefits and risks of non-pharmaceutical interventions (NPIs) during the pandemic.

## Methods

We undertook a systematic review to answer the question “What are the impacts of school closures on the health, wellbeing and educational outcomes of CYP?”, excluding impacts of closure on transmission of infection. Note we use ‘closure’ here to represent either full school closure or partial closure or dismissal (where schools remain open for small numbers of students).

The review was conducted according to PRISMA guidelines^15^ and was prospectively registered on the PROSPERO database (reference CRD42020181658).

### Search Strategy

We searched 11 electronic databases (PubMed, PsycINFO, Web of Science Social Citation Index, Australian Education Index, British Education Index, Education Resources Information Centre, WHO Global Research Database on COVID-19, Medrxiv, PsyArXiv, Research Square and COVID-19 Living Evidence) from inception to 1st September 2020. We used a combination of free text and Mesh terms related to children AND school AND school closure/social distancing measures (outlined in eTable 1). We screened the reference list of included articles and asked experts in the field for additional reports.

Inclusion criteria

1. Participants: any CYP aged 0-20 years.
2. Exposure: Nursery, preschool, primary or secondary school closure for any length of time in response to any non-routine event (e.g. pandemic, epidemic, disaster, weather, teacher strike, budget constraints) were included, whether implemented together with broader NPIs or alone. Literature on higher education were excluded as was literature on absences related to holidays, truancy or medical reasons
3. Comparators: For studies with control groups, the comparator was open schools or regions without lockdowns; for studies without control groups, the comparator was change from before closure.
4. Pre-specified outcomes: any physical or mental health and wellbeing outcome (eTable 2). We included health service use as a proxy measure of health outcome. We also included any available information on educational attainment and parent/ carer outcomes however these are published in a separate paper.
5. Types of studies: Observational quantitative studies including prospective and retrospective cohort studies; uncontrolled before after studies; modelling studies and cross sectional studies (included if provided information that could be compared with pre-lockdown normative data).
6. Publication status: published or pre-print studies

We adopted a machine learning (ML) approach^16^ for screening titles and abstracts, developed by the EPPI-Centre at the UCL Institute of Education and using EPPI-Reviewer 4 software.^17^ The ML algorithm was trained on the first 1500 articles and then a classifier model built to rank subsequent studies and identify a threshold below which studies were highly likely to not be relevant. Two researchers independently screened identified records on title/abstract and potentially relevant studies were acquired in full-text and independently assessed for inclusion by 2 researchers (HC/ SR/ SH/ JP). Decisions about inclusion were independently reassessed by the senior authors (RV, SM).

### Data Extraction and Quality Assessment

Two review authors (SM, RS) independently extracted outcome data from the studies, which were checked independently by a third (RV). Evidence was ranked by type of study and quality, which was independently rated by two authors (SM, RS), using: the Newcastle-Ottawa Scale (NOS)-Cohort studies for prospective and retrospective cohort studies;^18^ modified NOS for cross sectional studies;^19^ NHBLI tool for pre-post studies;^20^ and a modified checklist for modelling studies.^21^ Across checklists, studies were categorized as high quality if they met ≥90% of criteria, medium if ≥ 50% but <90%, and low if <50%.

### Data Synthesis and analyses

Due to heterogeneity of designs and measures, statistical meta-analysis was not possible. Instead we performed a narrative synthesis of the results, grouping studies according to the type of outcome then by study design, weighting interpretation by study type and quality. We aimed to undertake sub-analyses to examine: a) whether outcomes differed by exposure (differing causes of school closure (e.g. COVID-19 or not; school closure alone compared with school closure together with broader social ‘lockdown’); b) whether outcomes differed by socioeconomic status; and c) differences in outcomes by age or school-type.

## Results

Figure 1 shows the search flow. A total of 16,817 records were retrieved after removing duplicates, of which 151 were reviewed in full text as potentially relevant, and 79 studies (reported in 80 publications) were finally included. In this paper, we report the results from the 72 studies (73 publications) reporting CYP health outcomes (see Table 1). Other outcomes will be reported separately.

**Figure 1.**
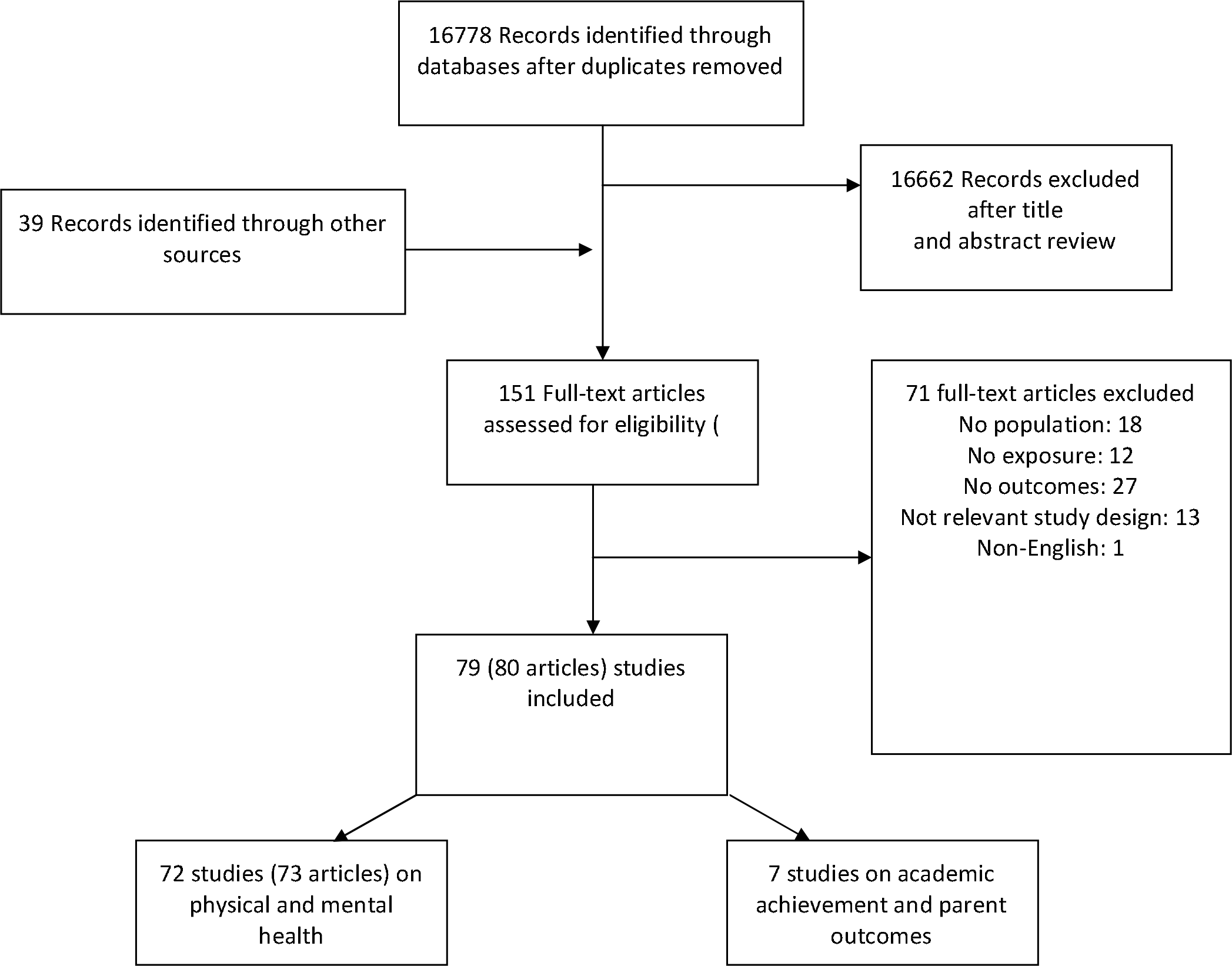
PRISMA 2009 Flow Diagram.

**Table 1.** Characteristics of included studies

| REFERENCE<br>(study, year) | Study<br>design | Period of reference | Country | Exposure | Reason for<br>school closure | School closure<br>/Lockdown<br>duration | Source of data | N subjects<br>enrolled | Mean Age<br>(range/ SD) | Male<br>% | SES<br>data |
| --- | --- | --- | --- | --- | --- | --- | --- | --- | --- | --- | --- |
| 1. AN 2020 | MS | From April 2020 to March 2021 | USA | SC | C-19P | **Scenario 1:** April-May 2020 nationwide SC. **Scen 2:** Scen 1 + 10% reduction in daily physical activity (June-August). **Scen 3:** Scen 2 + September - October SC. **Scen 4:** Scen 3 + November - December SC | Early Childhood Longitudinal Study, Kindergarten Class of 2010-2011 (ECLS-K:2011). | 15,631 children | 5-6 y up to 10-11 at the end of follow-up | NR | Y |
| 2. Angoulvant 2020 | TSA | From January 1, 2017 to April 19, 2020 | France | SC+L | C-19P | 1 m | Regional Center of Observation and Action on Emergencies e-CERVEAU (Agence Régionale de Santé) | 871,543 visits | NR | NR | N |
| 3. Baron 2020 | TSA | From January 2004 to March-April 2020 | USA | SC | C-19P | 1.5 m | Florida Department of Children and Families | 13.132 county-by-month observations | <18 y | NR | Y |
| 4. Baysun 2020 | UPP | Pre: March 2020; post: May 2020 | Turkey | SC+L | C-19P | 3 m | NR | 4 children | 14-30 m | 50% | N |
| 5. Bhopal 2020 | CHC | March 2020 compared to same period in 2018 - 2019 | UK | SC+L | C-19P | 1 m | Child protection medical examination services database | 107 children | 0-16 y | NR | N |
| 6. Bressan 2020 | CHC | January 1 - April 20, 2020 compared to same period 2019 | Italy | SC+L | C-19P | 2 m | PED electronic database University of Padua, | 3,713 visits | > 1 y | NR | N |
| 7. CDC 2010 | CSS | In 2009 | USA (39<br>States) | SC | IA (H1N1) | Up to 3 days<br>(58%);<br>up ≥ 5 days (26%) | TI | 523 households | <18 y | NR | N |
| 8. Chaiyachati 2020 | CHC | March 23- April 21 2020<br>compared to same period of<br>2017-2020 | USA | SC+L | C-19P | 1 m | PED electronic medical<br>record of the Children's<br>Hospital of Philadelphia. | 29,496 visits | 1-21 y | 52% | N |
| 9. Chandir<br>2020 | CHC | September 23, 2019- March<br>22, 2020 compared to March<br>23- May 9, 2020 | Pakistan | L | C-19P | NA | Electronic Immunization<br>Registry (Zindagi Mehfooz<br>Program; ZM EIR) | 701.324 children | 0-23 m | NR | Y |
| 10. Chen 2020 | PCS | From February 22, 2020 to<br>March 8, 2020 | China | SC+L | C-19P | 1 m | online survey | 7.772 Students | 12-18 y | 47.7% | N |
| 11. Children's<br>Society<br>2020 GL | CSS | In April - June 2020 | UK | SC+L | C-19P | 2 m | Online survey | Over 2.000<br>household and<br>150 children | 10-17 y | NR | Y |
| 12. Christoforidis 2020 | UPP | Pre: 3-weeks before March<br>10, 2020; post: 3-weeks after<br>March 10, 2020 | Greece | L | C-19P | 3 weeks | CareLink System data | 34 children or<br>caregivers | 2.5-18.5 y | 47.6% | N |
| 13. Ciofi Degli<br>Atti 2020 | CHC | (1) Jan 1 - Feb 19, 2020; (2)<br>Feb 20-March 10, 2020; and<br>(3) March 11- April 20, 2020. | Italy | SC+L | C-19P | 2 m | PED registry , Bambino<br>Gesù Children's Hospital | 18.825 visits | 1- >15 y | NR | N |
| 14. Cozzi 2020 | CHC | March 9 -April 13, 2020,<br>compared to Feb 2 -March 8<br>2020 | Italy | SC+L | C-19P | 1 m | PED database, Institute for<br>Maternal and Child Health<br>IRCCS | 3.362 visits | 6 y(range 2-11) | 51% | N |
| 15. Darlington<br>2020 | CSS | In April 6 - May 4, 2020 p | UK | L | C-19P | 1 m | Online survey | 171 parents | 7y median.<br>(range 1-24) | NR | Y |
| 16. Dayal 2020 | CHC | April 2020 compared to April 2019- March 2020 | India | L | C-19P | NR | Tertiary care paediatric referral hospital database | NR | NR | NR | N |
| 17. Della<br>Giulia<br>2020 | UPP | Feb 25 - March 25 2020 | Italy | SC+L | C-19P | 1 m | Questionnaire | 37 mothers | 3.81 y<br>(range 3–6) | 51% | N |
| 18. Di Giorgio<br>2020 | CSS | In April 1-9, 2020 p | Italy | SC+L | C-19P | 1 m | Online survey | 245 parents | 4.10 y (SD<br>0.92) | 53% | N |
| 19. Duan 2020 | CSS | NR | China | SC+L | C-19P | NR | Online survey | 3.613 children<br>and adolescents | 7-18 y | 50.2% | N |
| 20. Dunton<br>2020 | CSS | In April 25–May 16, 2020 | USA | SC+L | C-19P | 1-2 m | Online survey | 211 parents | 8.73 y (SD<br>2.58) | 47% | Y |
| 21. Dyson<br>2020 | CHC | March 23 -May 23, 2020<br>compared to same period<br>2019 | UK | SC+L | C-19P | 6 w | PED neurosurgery<br>electronic patient record<br>system. | 146 referrals | 5.63 y (SD<br>5.66) (pre C-19P); 4.84 y<br>(SD 4.91)<br>(during C—<br>19P) | 56.2% | N |
| 22. Effler 2010 | CSS | In 2009 | Australia | SC, class<br>cancelled | IA (H1N1) | 1 w | written questionnaire | 233 households | 11 y median<br>(range 5-13) | NR | N |
| 23. Ellis 2020 | CSS | In April 1, 2020 | Canada | SC+L | C-19P | 3 w | Online survey | 1.316 adolescents | 16.7 y | 21.9% | N |
| 24. Falkingham 2020 | CSS | In April 2020 | UK | SC+L | C-19P | 1 m | Understanding Society<br>COVID-19 Study | 895 adolescents | 16-24 y | NR | Y |
| 25. Gallagher 2020 | UPP | April 10 -May 22, 2020<br>baseline survey; 1 month<br>later 1st follow up | UK | SC+L | C-19P | 3 m | Online survey | 194 parents and<br>58 adolescents | 11-18 y | 9.30% | Y |
| 26. Garcia de<br>Avila 2020 | CSS | In April-May 2020 | Brazil | SC+L | C-19P | 1-2 m | Online survey | 289 children | 8.8 y | 45.7 y | Y |
| 27. Garstang 2020 | CHC | Late February- late June 2020<br>compared to same period<br>2018, 2019 | UK | SC+L | C-19P | 3 m | Electronic patient records<br>from Child Protection<br>Medical Examination<br>database | 200 referrals | 69 m<br>median (IQR<br>85) | 63.5% | N |
| 28. Garude 2020 | CHC | March 23, 2020- April 26<br>2020 compared to same<br>period 2019 | UK | SC+L | C-19P | 1 m | tertiary trauma centre<br>database. | 37 referrals | nr | NR | N |
| 29. Gelardi 2020 | CSS | In May 10 2020 | Italy | SC+L | C-19P | 2 m | telephonic interview | 120 parents | 5.69 y<br>(range 3-13) | NR | N |
| 30. Gift 2010 | CSS | In 2009 | USA | SC+L | IP (H1N1) | 1 w | telephone interview | 214 households<br>(269 students) | < 18 y | NR | Y |
| 31. Heymann 2004 and<br>2009 | UPP | *Heymann 2004*- (1) Pre:<br>January 4–17, 2000; (2)<br>During SC: January<br>18–31, 2000; (3) Post:<br>February 1–14, 2000<br>*Heymann 2009*: 5 December<br>to 25 March from 1998 to<br>2002 | Israel | SC | influenza<br>outbreak | 2 w | Administrative health<br>service database<br>(computerised data of the<br>Maccaby Health Service) | 186.094 children | 6-12 y | NR | N |
| 32. Iozzi 2020 | CHC | March 10 - May 3, 2020<br>compared to same period<br>2019. | Italy | SC+L | C-19P | 2 m | Patient records of PED San<br>Matteo Hospital,<br>University, Pavia, | 2.956 visits | NR | 53.6% | N |
| 33. Isumi 2020 | CHC | March-May 2020, compared<br>to same period in 2018 -2019 | Japan | SC+L | C-19P | 2 m | Public data on suicide<br>statistics, Ministry of<br>Health, Labour and Welfare | NR | < 20 y | NR | N |
| 34. Yeasmin<br>2020 | CSS | In April- May 2020 | Bangladesh | SC+L | C-19P | 1 m | Online survey | 384 parents | 5-15 y | NR | Y |
| 35. Johnson<br>2008 | CSS | In 2006 | USA | SC | influenza B | 10 | telephone interview by<br>questionnaire | 220 households<br>(355 students) | 12 median<br>(range 5–<br>19) | 50% | N |
| 36. Qi 2020 | CSS | In February 2020 | China | SC+L | C-19P | 1 w | Online survey | 9.954 adolescents | 11-20 y | NR | N |
| 37. Kilincel<br>2020 | CSS | NR | Turkey | SC+L | C-19P | nr | Online survey | 745 adolescents | 16.83 y (SD<br>1.66) | 30.5 | Y |
| 38. Krivec<br>2020 | CHC | March 16- April 20, 2020<br>compared to same period<br>previous 3 years | Slovenia | SC+L | C-19P | 1 m | Administrative hospital<br>data | NR | NR | NR | N |
| 39. Kuitunen<br>2020 | CHC | February 17-March 15, 2020<br>compared to March 16-April<br>12, 2020 | Finland | SC+L | C-19P | 1 m | Patient records of two PEDs<br>and Finnish national<br>Infectious Disease Register | 816 visits | NR | NR | N |
| 40. Lazzerini<br>2020 | CHC | Pre: March 2018- March<br>2019; Post: March 2020 | Italy | SC+L | C-19P | 1 m | Italian Paediatric Hospital<br>Research Network | 19.644 visits; 12<br>delayed<br>admission | NR | NR | N |
| 41. Levita<br>2020 | CSS | In March 23-28, 2020 and<br>April 22- May 1, 2020 | UK | SC+L | C-19P | 1 m | Online survey | 546 parents | 13-18 y | NR | N |
| 42. Lopez-Bueno<br>2020 | CSS | In 22 March - 10 May 2020 | Spain | SC+L | C-19P | 2 m | Online survey | 860 parents | 9.6 y (SD<br>3.9) | 50.8% | Y |
| 43. Lynn 2020 | CSS | In April 1, 2020 | UK | L | C-19P | 1 m | Online survey | 4.075<br>paediatricians | NR | NR | N |
| 44. Mann<br>2020 | CHC | March 23– May 31 2020<br>compared to same period<br>2019 | UK | SC+L | C-19P | 2.5 m | Emergency Care Data Set | 148 discharge<br>diagnosis | 0-18 y | NR | N |
| 45. Manzoni<br>2020 | CHC | March-April 2020 compared<br>to same period 2019 | Italy | SC+L | C-19P | 2 m | PED database of two<br>tertiary centres | 1.654 visits | 0-14y | NR | N |
| 46. Marino<br>2020 | CSS | April 9,2020 and May 9, 2020 | UK | L | C-19P | 2 m | Online survey | 184 parents and<br>36 young people | Children: 8 y<br>median (IQR<br>3-13);<br>young: 18 y<br>median (IQR<br>18-22) | NR | Y |
| 47. Martinelli<br>2020 | UPP | Post: March 8- April 20, 2020;<br>Pre: previous 8 weeks | Italy | SC+L | C-19P | 1.5 m | Italian regional pediatric<br>IBD referral center<br>database; Interviews | 180 parents and<br>children | 15 y (range<br>2-18) | 54% | N |
| 48. McDonald<br>2020 | CHC | January-April 2020 compared<br>to same period 2019 | UK | L | C-19P | NR | Electronic patient records<br>entered in SystmOne | 136.698 exavalent<br>vaccination;<br>127.173 MMR1<br>vaccination | NR | NR | N |
| 49. Nastro<br>2020 | UPP | Pre: March 9, 2020 - Post:<br>April 20, 2020; | Italy | SC+L | C-19P | 1 m | Online survey | 71 Parent or<br>adolescents | 1-18 y | NR | N |
| 50. Odd 2020 | CHC | January 1-March 23, 2020<br>compared to March 24- May<br>17, 2020. April 1- May 17,<br>2020 compared to same<br>period 2019 | UK | SC+L | C-19P | 2 m | National Child Mortality<br>Database (NCMD) | 51 children | <18 y | 58% | Y |
| 51. Ougrin<br>2020 | CHC | March -April 2020 compared<br>with same period 2017-2019 | UK | SC+L | C-19P | 2 m | National Commissioning<br>Data Repository and<br>NHS Digital data | 3.141 inpatient<br>admissions | 0-17 y | NR | N |
| 52. Pearcey<br>2020 | UPP | In April 17- June 22, 2020<br>baseline survey; 1st follow<br>up: 1 month later | UK | SC+L | C-19P | NR | Online survey | 972 parents | 2-5 y | NR | Y |
| 53. Pearcey<br>2020 B | UPP | In March 30- May 31, 2020<br>baseline survey; 1st follow<br>up: 1 month later | UK | SC+L | C-19P | NR | Online survey | 2.890 parents and<br>572 adolescents | 13 y | NR | Y |
| 54. Pietrobelli<br>2020 | UPP | Pre: May- June 2019; Post:<br>March 2020 | Italy | TSC+L | C-19P | 3 w | telephone interview | 41 Parents | 13.0 y<br>(range, 6-18) | 53.6% | N |
| 55. Roland<br>2020 | CSS | In April- May 2020 | UK | L | C-19P | 1 m | Online survey | 1.349<br>paediatricians | NR | NR | N |
| 56. Rose 2020 | CHC | March 21-April 26, 2020<br>compared to same period in<br>2019. | UK | SC+L | C-19P | 1.5 m | electronic health records of<br>single central PED, London | 4690 visits | 3 y (range 1-9) | 55% | N |
| 57. Roy 2020 | CSS | NR | India | SC+L | C-19P | NR | Online survey | 1.065 adolescents | 19.9 y (SD 3.5) | NR | N |
| 58. Russel 2020 | CSS | In April 27–28, 2020 | USA | SC+L | C-19P | 1 m | Online survey | 420 Caregivers | 0-18 y | NR | Y |
| 59. Segre 2020 | CSS | In May 18- June 7, 2020 | Italy | SC+L | C-19P | 3 m | Survey via video-meeting platform | 82 children and adolescents | 10.4 y (range 6-14) | 53.7% | Y |
| 60. Sheridan 2020 | CHC | April - April 29, 2020 compared to same period 2019 | USA | SC+L | C-19P | 1 m | Electronic medical system database of one tertiary care children's hospital | NR | NR | NR | N |
| 61. Sidpra 2020 | CHC | March 23 -April 23 2020 compared to same period in the previous 3 years | UK | SC+L | C-19P | 1.5 m | Administrative hospital data | 10 in 2020; NR in the previous years | 6 m | 60% | Y |
| 62. Timperio 2009 | CSS | In 2008 | USA | SC | SI | 3-4 d | telephone survey | 261 households (480 children) | NR | NR | Y |
| 63. Tittel 2020 | TSA | From 13 March to 13 May in each year (2011 -2020) | Germany | L | C-19P | 2 m | Diabetes Prospective Follow-up registry | 4.628 children and adolescents | 6 m-18 y | NR | N |
| 64. Tsai 2017 | CSS | In April 25-May 6, 2013 | US | SC | preparation for potential flooding | 8 d | Self-administered questionnaire | 208 (27%) households | NR | NR | Y |
| 65. Watson 2020 (A-D) | CSS | In June 22 - July 6, 2020 | UK | SC+L | C-19P | 2.5 m | Online survey | 11.228 Parents | 2–7-y | 50% | N |
| 66. Widnall<br>2020 | UPP | April/May 2020; compared to<br>pre-pandemic survey in<br>October 2019 | UK | SC+L | C-19P | 1.5 m | Online survey | 721-770<br>adolescents | 13-14 y | NR | N |
| 67. Xie 2020 | CSS | In End of February- Early of<br>March 2020 | China | SC | C-19P | 1 m | Online survey | 1784 children | 7-12 y | 56.7% | N |
| 68. Zheng<br>2020 | CSS | In February 2020 | China | SC+L | C-19P | 1 m | Online survey | 1.620 children | 10.10 y (SD<br>1.63) | 52.2% | N |
| 69. Zheteyeva,<br>Y. 2017 | CSS | In August 2012 | USA | SC | Preparation for<br>Hurricane Isaac | 4 d | Self administered<br>questionnaire | 2.229 household<br>(4.247 adults,<br>4.171 children) | 5-18 y | 48.4% | Y |
| 70. Zhou 2020<br>a | CSS | In February 2020 | China | SC+L | C-19P | 1 w | Online survey | 4.085 adolescents | 15 y (range:<br>11-18) | 0% | N |
| 71. Zhou 2020<br>b | CSS | In March 2020 | China | SC+L | C-19P | 3 w | Online survey | 8.079 adolescents | 16 y | 46.5% | N |
| 72. Zhou 2020<br>c | CSS | In March 2020 | China | SC+L | C-19P | 3 w | Online survey | 7.736 adolescents | 12-18 y | 46.5% | N |

Of the 72 studies, 24(33%) were cohort studies, 23 of which used an external historical/retrospective comparison group^22-44^ and 1 a parallel comparison group;^45^ 33 (46%) were cross-sectional studies;^46-78^ 14(19%) were uncontrolled pre-post studies, 11 of which included one measurement pre- and post-exposure^79-90^ and 3 included repeated measures; ^91-93^ and 1(1%) was a modelling study.^94^

Forty five studies (63%) were high-quality (eTables 3 to 7); 23 cohort studies,^22-44^ 17 cross-sectional,^46,48,52,54,56,59,61,62,64,65,67,71-74,76,78^ 4 pre-post,^82,83,91-93^ and 1 modelling study.^94^ Eighteen (25%) were medium-quality; 13 cross sectional,^49-51,53,55,57,58,60,66,68,70,75,77^ and 5 pre-post studies.^85,86,88-90^ Nine (13%) were low-quality; 1 cohort,^45^ 3 cross sectional,^47,63,69^ and 5 pre-post studies^79-81,84,87^

Eight were from China,^45,50,68,70,72-74,77^ 7 from other low and middle-income countries (LMIC; Turkey,^58,79^ Pakistan,^25^ India,^28,65^ Brazil,^55^ Bangladesh^76^) and 13 from Italy,^23,26,27,32,35,38,49,56,78,81,84-86^ 21 from the UK,^22,29-31,37,39-42,44,47,48,54,60,62-64,69,87,89,90^ 12 from the USA,^24,43,46,51,57,59,66,67,71,75,92,94^ and 1 study each from Ireland,^88^ France,^91^ Germany,^93^ Greece,^80^ Spain,^61^ Finland,^34^ Slovenia,^33^ Israel,^82,83^ Australia,^52^ Canada^53^and Japan.^36^

The exposure in all studies was the COVID-19 pandemic, with the exception of 5 studies (7%) of influenza outbreaks^46,52,57,59,66^ and 1 following a teacher strike.^82,83^

### PreCOVID-19 studies

Outcome data were confined to social activity and healthcare use. One cohort study from Israel reported that a 12-day teacher strike^82,83^ resulted in a relative risk of diagnosis of respiratory infection of 0.76 (95% CI 0.75– 0.77), and reductions in physician visits by 28%, emergency department (ED) attendances by 28% and medication purchases by 35% but no change in hospital admissions.^82,83^ Five cross-sectional surveys from the USA and Australia examined activities during brief school closures due to influenza outbreaks, and reported that 40-89% of CYP participated in activities outside the home or in public places,^46,52,57,59,66^

### COVID-19 studies

All reported changes relate to lockdown periods during the first pandemic wave (February/March to May/June 2020). Changes reported below refer to comparisons with either historical control periods in cohort studies, data collection prior to lockdown in pre-post studies, or, for cross-sectional studies, either comparison with historical reference data or retrospective recall of the period before lockdown (see Table 1).

### Healthcare use (19 studies; eTable 7)

Fifteen high-quality studies (14 cohort; 1 pre-post study) reported change in healthcare use in single hospitals/regional centres during the first COVID-19 wave compared with historical control periods.^23,24,26-29,31,33-35,37,38,42,44,91^ Four high-quality studies reported national data; one Italian cohort study of ED attendances,^32^ one German pre-post study of diabetes presentations^93^ and two UK cross-sectional studies of delayed presentations.^62,64^

Reductions in Emergency Department (ED) attendances by CYP were consistently high across all countries with estimates from 64% in Finland,^34^ 67% in the USA,^24^ 68% in France,^91^ and 67-84% in Italy,^23,26,27,32,35,38^ to 89.3% in one UK study.^42^ Attendances were reduced across most presentations, including fever and respiratory infections,^24,26,27,38,91^ trauma and injuries^24,26,27,38^ and burns,^37^ although injuries increased as a proportion of all cases.^35,38^ One Italian study provided data on the impact of school closures separately to full lockdown. They reported that ED attendances fell 24.6% in 2.5 weeks before lockdown, during which schools closed for the final week, before falling 66.7% during full lockdown from 11 March.^26^ Two HQ studies from the UK^42^ and the US^24^ reported that reductions in ED attendances were mainly for less acute presentations, whilst one Italian study reported a small non-significant rise in acute presentations.^27^

Reductions in hospital admissions in COVID-19 studies ranged from 31%-75% in Italy,^26,27,38^ 45.0% in France,^91^ 45-60% in Finland,^34^ and 68% in the USA^24^ to 85.7% in the UK.^42^ Another Italian study reported that admissions fell 9.5% in the weeks preceding lockdown (when schools closed) and 30.7% during full lockdown.^26^ The proportions of ED presentations admitted during lockdown rose 13.2% in the UK^42^ and 15.8% in USA^24^ for ward admissions and 5.8% (1.9, 10.0) for paediatric intensive care in the UK,^42^ with an Italian study reporting a 164% rise in the proportion of ED presentations admitted.^27^ Studies from a number of countries reported large reductions in admissions for fever and respiratory infection^24,26,27,33-35^ and for asthma (75.9%).^33^ Data on injury admissions were mixed, with Italian studies reporting from a 32%^27^ reduction to no change.^26^ One Italian study reported a five-fold increase in admissions for domestic accidents (IRR 5.0 (1.7, 14.6))^23^ while a second Italian study reported no increase.^26^ Two single-centre UK studies examined head trauma; one reported a very large (1493%) increase in suspected abusive head trauma^44^ while another reported only a moderate non-significant increase in head trauma of all types and no change in non-trauma neurosurgical referrals.^29^ A UK study reported a 80.6% reduction in hand trauma admissions.^31^ Two studies examined diabetes: a Indian study across 4 states reported a 79% reduction in diabetes admissions including a 75% decrease in presentations of new cases of diabetes during lockdown, with all new cases presenting with severe diabetic ketoacidosis.^28^ In contrast, a national German study found no impact of the pandemic on the incidence of new cases of type 1 diabetes.^93^

Four high-quality studies reported data on delayed presentations. A cross-sectional study of all paediatricians in the UK during the first month of lockdown found 32% of those in urgent care and 18% of other paediatricians had witnessed delayed presentations in the past fortnight, with 9 deaths considered to have resulted from the delay.^62^ Cohort data from 5 Italian children’s hospitals early in lockdown identified 12 serious delayed presentations over the previous week of whom 6 required intensive care and 4 died.^32^ In both studies, the main delayed presentations were diabetic ketoacidosis, sepsis and malignancy.^32,62^ All cases of severe ketoacidosis identified in a 4-state Indian cohort study represented delayed presentations.^28^ In contrast a cross-sectional UK study of 7 paediatric ED units in the second month of lockdown found that only 3.8% (51 CYP) of ED attendances were identified as having a delayed presentation, 6 of whom were admitted to hospital (1 to intensive care); delays were identified as largely due to parental reluctance to attend hospital.^64^

### Routine vaccinations (2 studies: eTable 8)

Two high-quality cohort studies used routine administrative data to examine impacts on CYP vaccination rates. A study from Karachi, Pakistan, found a reduction of 52.8% in daily infant immunisation visits early in lockdown, although this improved to 27.2% reduction by the end of lockdown.^25^ An English study found that first infant doses of hexavalent vaccine (against diphtheria, tetanus, pertussis, polio, Haemophilus influenzae type b and hepatitis B) early in lockdown had changed little compared with 2019, although there had been a 24.2% reduction in first measles-mumps-rubella vaccination. However, by the mid-point of lockdown, vaccination coverage for both vaccines was higher than in 2019.^39^

### Mental health and wellbeing (27 studies: eTable 9)

Studies originated from Japan,^36^ the UK,^40,41,47,60,69,87,89,90^ the USA,^43,75^ China,^45,50,68,70,72,74,77^ Italy,^49,78,85^ Turkey,^58^ Ireland,^88^ India,^65^ Canada,^53^ Brazil^55^ and Bangladesh.^76^ All studies conducted during lockdown were online or by telephone. Six (3 cohort, 3 cross-sectional) studies reported broadly representative data.^36,40,41,47,60,87^ The remainder were either longitudinal studies making use of pre-pandemic data collection for comparison or cross-sectional convenience samples which compared findings to pre-pandemic reference data.

#### Suicide

Two high-quality cohort studies found non-significant increases in national suicide rates compared with historical control periods in England (relative risk (RR) for <18 years: 1.41 (95% CI: 0.80, 2.46)^40^ and Japan (<20 years: incidence rate ratio (IRR) = 1.15 (0.81, 1.64)).^36^ In the English sample, factors related to the pandemic and lockdown were judged to contribute to 48% of deaths during lockdown.^40^

#### Mental health presentations

A high-quality national English cohort study found psychiatric inpatient admissions decreased by 40.2%, with large decreases in ED presentations for mental health reasons including self-harm.^41^ A high-quality US regional cohort study reported decreases in ED mental health presentations of just over 50%, with self-harm presentations reduced by 65.2%.^43^

#### Mental health symptoms

##### Representative surveys

A high-quality cross-sectional population-based study of UK young people during lockdown found that 53.3% of girls and 44.0% of boys aged 13-18 years had symptoms of anxiety and trauma above population threshold, with 47.4% of girls and 59.6% of boys reporting anxiety, while depressive symptoms were reported in 19.4% of girls and 21.9% of boys.^60^

A low-quality pre-post survey of young people in South-West England found reductions in proportions with anxiety symptoms in both boys and girls compared with October 2019, noting that mean anxiety scores fell amongst those with high scores pre-lockdown but there was little change in those with previously normal scores. Proportions with depressive symptoms rose slightly in girls and fell slightly in boys; however mean scores fell in those with pre-existing high scores but rose in those with previously normal scores.^87^ There were increases in young people’s sense of connection with school but no change in peer or family connection scores. Those with low pre-pandemic school connection showed greater reduction in anxiety scores but little change in depression scores. Anxiety and depression scores increased most in those with poorer connection with family and peers pre-pandemic.^87^

##### Convenience samples

A series of large cross-sectional surveys of mixed quality in Chinese school-aged CYP and a high-quality cohort study^45^ consistently found high levels of symptoms reaching clinical thresholds on self-report screening tools, and higher than recent reference data. Estimates for significant anxiety ranged from 10-19%^45,70,74,77^ Estimates for depressive symptoms ranged from 17-39% ^50,68,72,74^ although one study reported only 6.3%.^70^ One cohort^45^ and one-cross-sectional^68^ study found symptoms greater in Wuhan than other cities, consistent with greater exposure to lockdown.

Similar findings of higher proportions with problems than in reference populations were seen in large cross-sectional studies from other countries. Depressive symptoms were reported in 28% of Canadian young people^53^ and 26.5% of children from Bangladesh.^76^ Anxiety symptoms were reported in 19.4-21.8% of Brazilian children.^55^ Suicidal ideation was only reported in one medium-quality cross-sectional Canadian study, which found that 17.5% of 16-18 year olds reported suicidal ideation in the past week, compared to 6% in pre-pandemic estimates.^53^

A large low-quality Scottish cross-sectional study of 2-7 year olds found that 47% of parents reported worsening of their child’s behaviour and 45% reported worsening of their child’s mood, with proportions of 4-7 year olds with borderline or high scores for emotional difficulties (37%), conduct problems (43%) and hyperactivity/inattention (41%) approximately double that expected.^69^

Smaller cross-sectional studies of mixed quality from a range of countries reported high levels of stress,^75^ anxiety, ^49^ behavioural difficulties^49^ and hyperactivity/inattention^49^ in younger children and stress,^65^ anxiety^58^ and depressive symptoms in adolescents.^78^ Problems appeared greatest in those with previous mental health problems^58^ and where parents had poorer mental health.^75^

A low-quality cross-sectional consultation with 150 young people in England suggested 84% believed they had coped well overall during lockdown, with 70% reporting they coped well with schools closing. However 37% and 30% respectively reported coping poorly with not seeing friends and family.^47^

Change in psychological function during lockdown was examined by three medium-quality pre-post studies using national convenience samples to examine change over 1 month during lockdown. A study of 2-5 year old children in the UK found no changes in emotional difficulties but that restlessness/inattention difficulties reduced, whilst behavioural difficulties reduced significantly in boys but not girls during lockdown.^90^ Amongst primary school children (4-10 or 11 years), an Irish study found no significant change in mean scores for emotional, behavioural or restlessness/inattention difficulties,^88^ whilst a UK study found that emotional, behavioural and restlessness/inattention difficulties increased significantly.^89^ Amongst adolescents, the Irish study found no significant change in mean scores for emotional or behavioural difficulties amongst 12-18 year olds by either parental or adolescent report.^88^ In contrast, the UK study found significant increases in restlessness/inattention difficulties and decreases in emotional difficulties by parent report in 11-16 year olds, although young people themselves reported no change in difficulties.^89^ Parents of CYP with pre-existing mental health problems reported a significant reduction in their child’s emotional difficulties during lockdown in both the Irish and UK studies.^88,89^

#### Wellbeing

A large broadly-representative but low-quality cross-sectional study of 10-17 year olds in England during lockdown found low life-satisfaction in 18%, higher than in previous years (10-13%), and that low wellbeing scores, representing likely clinical problems, were found in 26.9% of 13-17 year olds.^47^ A broadly-representative low-quality pre-post survey from South-West England reported minor worsening of mean wellbeing scores from before to during lockdown, changes the authors regarded as not meaningful.^87^

##### Child abuse (3 studies: eTable 10)

A high-quality time-series study from Florida estimated that the number of notifications of child abuse in the state of Florida decreased by 27% during lockdown, using school staffing and spending data to conclude this resulted from school closures.^92^ Two high-quality large regional cohort studies from the UK estimated that child protection medical referrals fell 36^22^-39%,^30^ with one estimating that the proportion of referrals originating from schools approximately halved.^30^

##### Sleep (10 studies; eTable 11)

A high-quality nationally-representative UK cohort study found that 25% of 16-24 year olds reported new onset of sleep problems due to worrying.^54^ A large low-quality cross-sectional convenience study of Scottish children 2-7 years found that proportions of children sleeping through the night (32% in 2-4 year olds and 50% for 5-7 year olds) were lower than pre-pandemic national data (38% and 60% respectively), with 33% of parents reporting worse sleep since the pandemic and only 7% sleeping better.^69^ No changes in sleep duration or quality were reported by cross-sectional convenience studies in Italian^49^ and Spanish^61^ children; however a small pre-post study of Italian preschool children found a decrease in sleep duration early during lockdown^81^ and a cross-sectional study in Italian children found 61% reported difficulties falling asleep and fragmented sleep.^78^

Two cross-sectional convenience studies of Chinese young people reported increased sleep problems; one study reported 63.9% slept for 8 hours or less per night^72^ whilst the second found a prevalence of symptoms of insomnia in 23.2%.^73^ In contrast, increased sleep duration during lockdown was reported by a cross-sectional study of 13-25 year olds in India^65^ and a small pre-post study of young people with obesity in Italy.^86^

### Health Behaviours (eTable 12)

#### Physical activity (PA) and sedentary behaviour

In cross-sectional convenience samples from the US, Scotland and India, 36-47%^51,65,69^ of CYP experienced falls in PA, whilst 24-24.4% undertook more PA,^65,69^ whilst a Spanish study reported that mean daily PA fell 52%.^61^ A medium-quality pre-post study in Italian CYP with obesity found a decrease in PA of 2.3 hours per week (64% relative decrease).^86^ A medium-quality cross-sectional convenience study found that 41% of parents reported their child had done much more sitting compared with recall of the period before lockdown.^51^

#### Screentime and social media

Two cross-sectional convenience studies reported increases in screentime, although studies did not separate recreation from online learning; a Spanish study found that mean daily screentime rose by 2.9 hours per day (245% increase), with the greatest rises amongst teenagers,^61^ whilst an Indian study found mean screentime was 5.1 hours during lockdown, over 70% higher than previous national data.^65^ A medium-quality pre-post study in Italian CYP with obesity found a significant increase of 4.9 hours per day (296% increase).^86^

Increases in social media use were reported in two studies. A Canadian medium-quality cross-sectional convenience study found the proportion of older teenagers using social media >3 hours per day more than doubled from 31.9% to 77.2%.^53^ A broadly representative low-quality pre-post study of adolescents in South-West England found an increase in weekday high social media use (≥3 hours per day) amongst girls (42% pre-pandemic, 55% lockdown) but not boys (29% pre, 30% lockdown) but no change during weekends.^87^

#### Eating and diet

An large low-quality cross-sectional convenience study of Scottish children aged 2-7 years found little evidence of change in diet;^69^ however, cross-sectional convenience studies from India, Spain and Italy suggested an increase in overall levels of consumption,^65,78^ particularly of unhealthy food,^65,78^ and a reduction in fruit and vegetable consumption.^61^ A medium-quality pre-post study in Italian CYP with obesity found an increase in the number of meals eaten per day (4.2 to 5.3), with increased intake of potato chips and sugary drinks.^86^ We identified no studies of eating behaviours.

#### Substance use

No studies provided data on use of tobacco, alcohol or other drugs.

### Overweight (eTable 13)

A high-quality US microsimulation study estimated that 2 months of school closure would result in a 11.1% rise in childhood obesity in young children over the following year, with larger rises if social distancing reduced PA or there were additional school closures over the following year.^94^ A low quality pre-post study from Turkey reported that weight centile increased in young children from 25-50% centile to 50-75% centile.^79^

### Impacts on pre-existing conditions (eTable 14)

Six small-scale studies used cross-sectional or pre-post designs to examine the impact of school closures and lockdown on CYP with pre-existing conditions. Three Italian studies found reductions in admissions or improvements in symptoms in conditions including adenotonsillar hypertrophy,^56^ inflammatory bowel disease (IBD),^84^ and coeliac disease,^85^ although a pre-post study of diabetes control found no change in blood glucose control, insulin dose or carbohydrate intake in children on insulin pumps.^80^

Two cross-sectional UK studies, of cancer^48^ and congenital heart disease,^63^ found widespread marked parental concerns about the safety of their CYP and that 70-85% of parents believed that hospitals were not safe places for their child, anxieties that were shared by their CYP. Whilst one LQ Italian study found that 25% of CYP with IBD suspended or delayed immunoregulatory treatment against medical advice,^84^ a HQ English study found that only 2.3% of parents had reduced the amount of cancer chemotherapy they gave their child.^48^

We identified no included articles on impacts on children with learning difficulties or autism.

### Impacts of socioeconomic status

Few studies considered how socioeconomic status modified outcomes. A US cohort study found larger falls in ED attendances amongst African-American patients and those with public insurance;^24^ a cohort study from Pakistan found greater declines in child vaccination in the poorest communities;^25^ and a cross-sectional study from Brazil found greater anxiety in CYP from families with lower education levels.^55^ Pre-post studies found the UK^87,89^ and Ireland^88^ found few differences in change in psychological function over a month in lockdown, although higher-income parents reported significant increases in children’s behaviour problems during lockdown whilst lower-income parents did not,^89^ and employed parents reported significant reductions in preschool children’s behavioural and restlessness/inattentional difficulties whilst unemployed parents did not.^90^

## Discussion

This is the first comprehensive systematic review of the effects of school closures on CYP health and wellbeing. In addition to providing education, schools have important roles in promoting child development, wellbeing and mental health, forming part of child protection surveillance systems and providing access to health goods such as vaccination and mental health services.^95^ We found that almost all of the 72 studies from 20 countries (8 LMIC) included here documented harms to CYP that occurred during school closures and social lockdown, the vast majority during the first wave of the COVID-19 pandemic.

The strength of evidence from included studies was mixed. Stronger evidence was provided by the one-third of studies which were cohort studies that used pre-pandemic comparison data on the same population and the one-fifth that were longitudinal studies following the same population from before to during lockdown. However 46% were cross-sectional studies which relied upon comparison with pre-pandemic population norms to identify change. Furthermore, few cross-sectional studies were truly representative of the populations studied, with the difficulties of data collection during lockdown meaning that most used convenience sampling with its inherent biases. Whilst findings from convenience samples must be treated with caution, in some areas they provided the only available data in CYP whilst adults were the subject of higher quality studies.

We identified few studies prior to the COVID-19 pandemic, and few studies allowed us to separate the impacts of school closures from broader social lockdown due to the implementation of school closures as part of broader measures in the first wave in most countries.^3^ Health service use was the only area where studies were informative in this regard: there were smaller reductions in physician, ED and hospital attendances after closures due to a teachers’ strike than were seen from COVID-19, and ED attendances fell less in Italy during COVID-19 school closures than later during full lockdown. This suggests that for health service use at least, school closures account for only some of the changes seen during COVID-19.

The largest number of studies (27) concerned mental health and wellbeing. Whilst the strength of evidence was mixed, with a smaller number of cohort and longitudinal studies and many cross-sectional studies, evidence for impacts upon mental health and wellbeing was substantial and consistent. The great majority of high- and medium-quality studies, including the only nationally representative survey,^60^ identified considerable impacts across the range of emotional, behavioural and restlessness/inattention problems and overall psychological wellbeing. Both representative and large convenience studies including studies from high-income and LMIC found that 18-60% of CYP scored above thresholds suggesting they were at risk for psychological distress, particularly anxiety and depressive symptoms. In most studies, these proportions were substantially higher than before the pandemic. Whilst convenience samples are likely to inflate estimates of distress, these findings were consistent across study types. Risk appeared highest where CYP or parents had pre-existing mental health problems. There were some data from China to suggest that impact was greater where lockdown was more severe or prolonged. Studies from England and Japan found non-significant rises in suicide rates, although numbers of deaths remained very low, and a cross-sectional Canadian study reported an increase in suicidal ideation amongst CYP. These findings are consistent with other systematic reviews of the impacts of isolation on CYP mental health,^96^ and with longitudinal studies from China^97^ and the UK^98^ published after our search. We found no data on impacts on CYP with learning disability or special educational needs, although we note that a study published after our search found that CYP with autism and attention-deficit hyperactivity disorder had greater elevation of problem scores.^99^

In contrast to these findings, one pre-post study of English young people found improvements in mean anxiety scores during lockdown, particularly in those with pre-existing high scores and in those with poorer relationships with school.^87^ Some young people find attending school stressful and it is likely that for some CYP, time at home with care-givers may have strengthened social support and the sense of cohesion in some families or communities.^100^

The second largest group of studies were on health service use, consisting predominantly of high quality cohort studies, many using routine administrative data. Health service use is a proxy for health status but also heavily influenced by access to healthcare, which may have been reduced during the pandemic. Studies from all countries showed markedly reduced ED presentations and hospital admissions, particularly for low risk presentations and those with fever and respiratory tract infections. Whilst there was evidence of harm to CYP from delayed presentations in studies early in the pandemic, evidence from high-quality studies from three countries suggested that high risk presentations were not reduced overall, indicating that health systems in high-income countries functioned to avoid harm to CYP. Evidence from LMIC is confined to a single HQ study of diabetes,^28^ and findings of harms from delayed presentations raise the possibility that harm may have accrued to CYP where health systems are less accessible and resilient. Few data were available on impacts upon CYP with pre-existing conditions. Data suggested there may be improvements in symptoms related to reductions in circulating respiratory viruses with evidence of high levels of anxiety amongst CYP with complex conditions and their parents. Data on vaccination rates were sparse but suggested potential widening of inequalities in vaccination coverage. No data were identified on vaccinations delivered in schools.

Despite the rises in psychological distress described above, presentations for self-harm and psychiatric admissions were markedly reduced in high-quality cohort studies from two countries. This suggests an escalation of unmet mental health need during lockdowns which may bring additional harms to already vulnerable CYP. A reduction in the ability of the health and social care systems to protect children is evidenced by the large falls in child protection referrals seen in high-quality cohort studies from the USA and UK, with a halving of the expected number of referrals originating in schools.

Data on impacts upon health behaviours were drawn predominantly from cross-sectional studies using convenience samples. There was consistent evidence from medium- and high-quality studies that time spent using screens, including for social media, increased markedly when schools were closed in high-income countries and LMIC. Whilst some of the increase in screentime may have reflected online learning, evidence from outside our review suggests that young people themselves are concerned about the impact of high amounts of screen time on their wellbeing.^101^

The evidence on sleep, PA and diet are mixed in both findings and in quality. All data were parent or self-report, which are known to have issues for dietary behaviours^102^ and physical activity.^103^ Data suggest that levels of PA decreased and increased in different groups of CYP, with most studies suggesting greater numbers suffered decreases. Data on diet provide no clear signal but sound a warning that school closures and lockdowns may have impacts on groups such as CYP with obesity. No studies within our time frame and search strategy examined the impact on alcohol, tobacco or other drug use. Studies reported both longer and shorter sleep duration for CYP during lockdown, although reports of poorer sleep quality were dominant. High-quality studies from the UK and China suggested around one-quarter of young people developed significant sleeping difficulties during lockdown.

We identified no population-based studies that measured change in eating behaviours and weight-status. Modelling data from the US suggest concerning potential rises in childhood obesity related to the loss of school-based PA. Data suggest those CYP already overweight and obese maybe particularly vulnerable to the health impacts of school closure/lockdown.

Lack of data on socioeconomic status meant we cannot draw firm conclusions on how poverty might moderate the indirect impacts of the pandemic. Available data suggested it is likely there will be higher impacts in those from more deprived populations, widening already existing inequalities.

### Strengths and limitations

We undertook a high quality systematic review across a large number of electronic databases, educational as well as health databases and including preprints, with independent checking of study eligibility, data extraction and quality assessment. Studies were included from 20 countries across all income levels, although few studies from very resource-poor settings were included. Our findings are subject to a number of limitations. The majority of studies provided relatively low quality evidence. Studies were largely unable to reach or recruit new participants during lockdown, hence the reliance on online self-report data collection from convenience studies. Many publications were preliminary reports or preprints, and included only simple analyses which did not take account of potential confounders. Many studies used historical control periods, which in some failed to take account of seasonal variation. Studies using parent report may have been biased by greater amounts of time spent by parents with their CYP compared with pre-pandemic. We identified no data on the impact of the degree of school closures; note that in some countries (e.g. the UK), whilst schools were essentially closed, approximately 5% of students were in school during the first pandemic wave. We could not identify nor include studies on a number of important outcomes or vulnerable groups, including studies of children with learning difficulties or autism or studies of eating disorders or substance use.

## Conclusions

School closures as part of broader social distancing measures are associated with considerable harms to CYP health and wellbeing, in addition to potential impacts upon learning and family outcomes not considered here. These harms occurred at a time when access to health and social care was very markedly reduced and at a time when CYP were much less visible to protective systems. All COVID-19 data included here are short-term and relate to the first pandemic wave. There is concerning evidence that harmful changes in PA, screentime and diet can continue once schools are reopened,^104^ emphasising the potential for persisting harms. Longer-term harms are likely to be magnified by further school closures in subsequent waves. Data on longer-term impacts using strong research designs are urgently needed, particularly amongst vulnerable groups. This will require investment in new data collection systems as the exclusion of school-aged CYP from most national and international data-collection is well documented.^105^ Our findings are important for policy-makers seeking to balance the risks of transmission through school-aged children with the harms of closing schools, and be useful to those tasked with mitigating the harms of this pandemic for the next generation.

## Data Availability

All included data in this systematic review are drawn from published or preprint articles that are freely available.

## Acknowledgements

We are grateful to members of the WHO Research Advisory Group on COVID in Education Settings for suggestions on the work. We are also grateful to Alexandra Jamieson (UCL) for helping with screening of abstracts.

## Contributions

The review was conceptualised by RV, SM, NS and AM. The protocol for the review was developed by HC, SR, RV and SM with input from NS and AM. Data searches were undertaken by SR, HC, SH and JP with advice from CS. Preprint searches were undertaken by RV. Data extraction and quality assessment was undertaken by SM, RS and RV. Synthesis of data and writing of the manuscript was led by RV and SM. All authors contributed to commenting on drafts the manuscript.

