## Supplemental Tables for "Impacts of school closures on physical and mental health of children and young people: a systematic review"

### Supplement

#### etable 1 . Search strategies

PubMed

((((child, preschool[mh]) OR (adolescent[mh]) OR (child[tiab] OR children[tiab] OR child’s[tiab] OR "young people"[tiab] OR "young person*"[tiab] OR youth*[tiab] OR infant*[tiab] OR schoolchild*[tiab] OR "school child*"[tiab] OR boy*[tiab] OR girl*[tiab] OR teen*[tiab] OR adolescen*[tiab] OR pediatric*[tiab] OR paediatric*[tiab] OR peadiatric*[tiab])))) AND (((((((schools[mh:noexp]) OR (schools, nursery[mh]) OR (nurseries, infant[mh]) OR (school*[tiab] OR preschool*[tiab] OR "pre school*"[tiab] OR pre-school*[tiab] OR nursey[tiab] OR nurseries[tiab] OR kindergarten*[tiab] OR kindergarden*[tiab]) OR (("day care"[tiab] OR daycare[tiab]) AND (child*[tiab] OR infant*[tiab])))) AND ((close[tiab] OR closed[tiab] OR closure[tiab] OR closures[tiab] OR closing[tiab])))) OR ((“education access”[tiab] OR “access to education”[tiab]) OR ((“primary education”[tiab] OR “secondary education”[tiab]) AND (“no access”[tiab] OR “restricted access”[tiab] OR restrict*[tiab])))) OR ((quarantine*[tiab] OR self-isolat*[tiab] OR "lock down*"[tiab] OR lockdown*[tiab] OR lock-down*[tiab] OR "class dismissal*"[tiab] OR "school dismissal*"[tiab] OR "social distanc*"[tiab] OR "stay at home" [tiab] OR "shut down*"[tiab] OR shut-down*[tiab] OR shutdown*[tiab] OR "staying at home"[tiab])))

| [#1](https://www.ncbi.nlm.nih.gov/pubmed/advanced) | Search ((child, preschool[mh]) OR (adolescent[mh]) OR (child[tiab] OR children[tiab] OR child’s[tiab] OR "young people"[tiab] OR "young person*"[tiab] OR youth*[tiab] OR infant*[tiab] OR schoolchild*[tiab] OR "school child*"[tiab] OR boy*[tiab] OR girl*[tiab] OR teen*[tiab] OR adolescen*[tiab] OR pediatric*[tiab] OR paediatric*[tiab] OR peadiatric*[tiab])) |
| --- | --- |
| [#2](https://www.ncbi.nlm.nih.gov/pubmed/advanced) | Search (((schools[mh:noexp]) OR (schools, nursery[mh]) OR (nurseries, infant[mh]) OR (school*[tiab] OR preschool*[tiab] OR "pre school*"[tiab] OR pre-school*[tiab] OR nursey[tiab] OR nurseries[tiab] OR kindergarten*[tiab] OR kindergarden*[tiab]) OR (("day care"[tiab] OR daycare[tiab]) AND (child*[tiab] OR infant*[tiab])))) AND ((close[tiab] OR closed[tiab] OR closure[tiab] OR closures[tiab] OR closing[tiab])) |
| [#3](https://www.ncbi.nlm.nih.gov/pubmed/advanced) | Search (“education access”[tiab] OR “access to education”[tiab]) OR ((“primary education”[tiab] OR “secondary education”[tiab]) AND (“no access”[tiab] OR “restricted access”[tiab] OR restrict*[tiab])) |
| [#4](https://www.ncbi.nlm.nih.gov/pubmed/advanced) | Search (quarantine*[tiab] OR self-isolat*[tiab] OR "lock down*"[tiab] OR lockdown*[tiab] OR lock-down*[tiab] OR "class dismissal*"[tiab] OR "school dismissal*"[tiab] OR "social distanc*"[tiab] OR "stay at home" [tiab] OR "shut down*"[tiab] OR shut-down*[tiab] OR shutdown*[tiab] OR "staying at home"[tiab]) |
| [#5](https://www.ncbi.nlm.nih.gov/pubmed/advanced) | Search #2 OR #3 OR #4 |
| [#6](https://www.ncbi.nlm.nih.gov/pubmed/advanced) | Search #1 AND #5 |

PsycInfo – Ovid SP

| 1 | (child* or "young people" or "young person*" or youth* or infant* or schoolchild* or "school child*" or boy* or girl* or teen* or adolescen* or pediatric* or paediatric* or peadiatric*).ti,ab. |
| --- | --- |
| 2 | schools/ or Junior High Schools/ or High Schools/ or Middle Schools/ or Nursery Schools/ or Elementary Schools/ |
| 3 | (school* or preschool* or "pre school*" or pre-school* or nursey or nurseries or kindergarten* or kindergarden* or (("day care" or daycare) and (child* or infant*))).ti,ab. |
| 4 | (close or closed or closure or closures or closing).ti,ab. |
| 5 | 2 or 3 *(4 or 6)* |
| 6 | 4 and 5 *(7 and 9)* |
| 7 | ("education access" or "access to education" or (("primary education" or "secondary education") and ("no access" or "restricted access" or restrict*))).ti,ab. |
| 8 | (quarantine* or self-isolat* or "lock down*" or lockdown* or lock-down* or "class dismissal*" or "school dismissal*" or "social distanc*" or "stay at home" or "shut down*" or shut-down* or shutdown* or "staying at home").ti,ab. |
| 9 | 6 or 7 or 8 *(10 or 12 or 14)* |
| 10 | 1 and 9 *(2 and 15)* |

Web of Science - Social Citation Index

TS=((child OR children OR child’s or "young people" or "young person*" or youth* or infant* or schoolchild* or "school child*" or boy* or girl* or teen* or adolescen* or pediatric* or paediatric* or peadiatric*) and (((school* or preschool* or "pre school*" or pre-school* or nursery or nurseries or kindergarten* or kindergarden* or (("day care" or daycare) and (child* or infant*))) and (close or closed or closure or closures or closing)) or ("education access" or "access to education" or (("primary education" or "secondary education") and ("no access" or "restricted access" or restrict*))) or (quarantine* or self-isolat* or "lock down*" or lockdown* or lock-down* or "class dismissal*" or "school dismissal*" or "social distanc*" or "stay at home" or "shut down*" or shut-down* or shutdown* or "staying at home")))

**(year to date 385)**

| # 1 | TS=(child OR children OR child’s OR “young people”} OR “young person*” OR youth* OR infant* OR schoolchild* OR “school child*” OR boy* OR girl* OR teen* OR adolescen* OR pediatric* OR paediatric* OR peadiatric*)  *Indexes=SSCI Timespan=All years* |
| --- | --- |
| # 2 | TS=((school* OR preschool* OR “pre school*” OR pre-school* OR nursery OR nurseries OR kindergarten* OR kindergarden* OR (("day care" OR daycare) AND (child* OR infant*))) AND (close OR closed OR closure OR closures OR closing))  *Indexes=SSCI Timespan=All years* |
| # 3 | TS=(“education access” OR “access to education” OR ((“primary education” OR “secondary education”) AND (“no access” OR “restricted access” OR restrict*)))  *Indexes=SSCI Timespan=All years* |
| # 4 | TS=(quarantine* OR self-isolat* OR “lock down*” OR lockdown* OR lock-down* OR “class dismissal*” OR “school dismissal*” OR “social distanc*” OR “stay at home” OR “shut down*” OR shut-down* OR shutdown* OR “staying at home”)  *Indexes=SSCI Timespan=All years* |
| # 5 | #4 OR #3 OR #2  *Indexes=SSCI Timespan=All years* |
| # 6 | #5 AND #1  *Indexes=SSCI Timespan=All years* |

British Education Index (EBSCO host)

| S1 | (MH "children") OR (MH "Adolescents") OR (TI (child OR children OR child’s OR "young people" OR "young person*" OR youth* OR infant* OR schoolchild* OR "school child*" OR "school*child*" OR boy* OR girl* OR teen* OR adolescen* OR Pediatric* OR Paediatric* OR Peadiatric*)) OR (AB (child OR children OR child’s OR "young people" OR "young person*" OR youth* OR infant* OR schoolchild* OR "school child*" OR "school*child*" OR boy* OR girl* OR teen* OR adolescen* OR Pediatric* OR Paediatric* OR OR Peadiatric*)) |
| --- | --- |
| S2 | (MH "Schools") OR (MH "Nursery schools") OR (MH "Primary education") OR (MH "Secondary education") OR (MH "Preschool education") OR (MH "High schools") OR(MH "Kindergarten") OR (TI (school* OR preschool* OR "pre school*" OR pre-school* OR nursery OR nurseries OR kindergarten* OR kindergarden*)) OR (TI (("day care" OR daycare) AND (child* OR infant*))) OR (AB (school* OR preschool* OR "pre school*" OR pre-school* OR nursery OR nurseries OR kindergarten* OR kindergarden*)) OR (AB (("day care" OR daycare) AND (child* OR infant*))) |
| S3 | (TI (close OR closed OR closure OR closures OR closing)) OR (AB (close OR closed OR closure OR closures OR closing)) |
| S4 | S2 AND S3 |
| S5 | (TI ("education access" OR "access to education")) OR (AB ("education access" OR "access to education")) OR ((TI ("primary education" OR "secondary education") AND ("no access" OR "restricted access" OR "restrict*"))) OR (AB ("primary education" OR "secondary education") AND ("no access" OR "restricted access" OR "restrict*")))) |
| S6 | (TI ( quarantine* OR self-isolat* OR "lock down*" OR lockdown* OR "lock-down*" OR "class dismissal" OR "school dismissal*" OR "social distanc*" OR "stay at home" OR "shut down*" OR "shut-down* " OR "shutdown*" OR "staying at home")) OR (AB (quarantine* OR self-isolat* OR "lock down*" OR lockdown* OR "lock-down*" OR "class dismissal" OR "school dismissal*" OR "social distanc*" OR "stay at home" OR "shut down*" OR "shut-down* " OR "shutdown*" OR "staying at home")) |
| S7 | S4 OR S5 OR S6 |
| S8 | S1 AND S7 |

Australian Education Index- ProQuest

(MAINSUBJECT.EXACT.EXPLODE("Children") OR MAINSUBJECT.EXACT.EXPLODE("Adolescents") OR TI,AB(child OR children OR child’s OR "young people" OR ("young person" OR "young persons") OR youth* OR infant* OR schoolchild* OR ("school child" OR "school childcare" OR "school children") OR "school*child*" OR boy* OR girl* OR teen* OR adolescen* OR Pediatric* OR Paediatric* OR Peadiatric*)) AND ((((MAINSUBJECT.EXACT.EXPLODE("Schools") OR MAINSUBJECT.EXACT.EXPLODE("Nursery schools") OR MAINSUBJECT.EXACT.EXPLODE("Primary schools") OR MAINSUBJECT.EXACT.EXPLODE("Primary education") OR MAINSUBJECT.EXACT.EXPLODE("Secondary education") OR MAINSUBJECT.EXACT.EXPLODE("Preschools") OR MAINSUBJECT.EXACT.EXPLODE("High schools") OR MAINSUBJECT.EXACT.EXPLODE("Kindergarten") OR TI,AB(school* OR preschool* OR ("pre school" OR "pre schooler" OR "pre schoolers" OR "pre schools") OR pre-school* OR nursery OR nurseries OR kindergarten* OR kindergarden*)) OR TI,AB(("day care" OR daycare) AND (child* OR infant*))) AND (MAINSUBJECT.EXACT.EXPLODE("School closing") OR TI,AB(close OR closed OR closure OR closures OR closing))) OR (TI,AB("education access" OR "access to education") OR (TI,AB("primary education" OR "secondary education") AND ("no access" OR "restricted access" OR "restrict*"))) OR TI,AB( quarantine* OR self-isolat* OR "lock down*" OR lockdown* OR "lock-down*" OR "class dismissal" OR "school dismissal*" OR "social distanc*" OR "stay at home" OR "shut down*" OR "shut-down* " OR "shutdown*" OR "staying at home"))

| **1** | [MAINSUBJECT.EXACT.EXPLODE("Children") OR MAINSUBJECT.EXACT.EXPLODE("Adolescents") OR TI,AB(child OR children OR child’s OR "young people" OR ("young person" OR "young persons") OR youth* OR infant* OR schoolchild* OR ("school child" OR "school childcare" OR "school children") OR "school*child*" OR boy* OR girl* OR teen* OR adolescen* OR Pediatric* OR Paediatric* OR Peadiatric*)](https://search-proquest-com.libproxy.ucl.ac.uk/recentsearches.recentsearchtabview.recentsearchesgridview.scrolledrecentsearchlist.checkdbssearchlink:rerunsearch/9332566957214585PQ/None?site=australianeducationindex&t:ac=RecentSearches) | Australian Education Index |
| --- | --- | --- |
| **2** | [(MAINSUBJECT.EXACT.EXPLODE("Schools") OR MAINSUBJECT.EXACT.EXPLODE("Nursery schools") OR MAINSUBJECT.EXACT.EXPLODE("Primary schools") OR MAINSUBJECT.EXACT.EXPLODE("Primary education") OR MAINSUBJECT.EXACT.EXPLODE("Secondary education") OR MAINSUBJECT.EXACT.EXPLODE("Preschools") OR MAINSUBJECT.EXACT.EXPLODE("High schools") OR MAINSUBJECT.EXACT.EXPLODE("Kindergarten") OR TI,AB(school* OR preschool* OR ("pre school" OR "pre schooler" OR "pre schoolers" OR "pre schools") OR pre-school* OR nursery OR nurseries OR kindergarten* OR kindergarden*)) OR TI,AB(("day care" OR daycare) AND (child* OR infant*))](https://search-proquest-com.libproxy.ucl.ac.uk/recentsearches.recentsearchtabview.recentsearchesgridview.scrolledrecentsearchlist.checkdbssearchlink:rerunsearch/5178F189C9AF4E83PQ/None?site=australianeducationindex&t:ac=RecentSearches) | Australian Education Index |
| **3** | [MAINSUBJECT.EXACT.EXPLODE("School closing") OR TI,AB(close OR closed OR closure OR closures OR closing)](https://search-proquest-com.libproxy.ucl.ac.uk/recentsearches.recentsearchtabview.recentsearchesgridview.scrolledrecentsearchlist.checkdbssearchlink:rerunsearch/B7E757FE2E324AADPQ/None?site=australianeducationindex&t:ac=RecentSearches) | Australian Education Index |
| **4** | 2 AND 3 | Australian Education Index |
| **5** | [TI,AB("education access" OR "access to education") OR (TI,AB("primary education" OR "secondary education") AND ("no access" OR "restricted access" OR "restrict*"))](https://search-proquest-com.libproxy.ucl.ac.uk/recentsearches.recentsearchtabview.recentsearchesgridview.scrolledrecentsearchlist.checkdbssearchlink:rerunsearch/CAF43A9DD19D4D15PQ/None?site=australianeducationindex&t:ac=RecentSearches) | Australian Education Index |
| **6** | [TI,AB( quarantine* OR self-isolat* OR "lock down*" OR lockdown* OR "lock-down*" OR "class dismissal" OR "school dismissal*" OR "social distanc*" OR "stay at home" OR "shut down*" OR "shut-down* " OR "shutdown*" OR "staying at home")](https://search-proquest-com.libproxy.ucl.ac.uk/recentsearches.recentsearchtabview.recentsearchesgridview.scrolledrecentsearchlist.checkdbssearchlink:rerunsearch/3A587A6818E147FEPQ/None?site=australianeducationindex&t:ac=RecentSearches) | Australian Education Index |
| **7** | 4 OR 5 OR 6 | Australian Education Index |
| **8** | 1 AND 7 | Australian Education Index |
| **9** | 8 NOT conference papers |  |

Educational Resources Information Center (EBSCO)

| S1 | DE("Children") OR DE("Adolescents") OR DE("Child health") OR (TI ("Child health" OR child OR children OR child’s OR "young people" OR "young person*" OR youth* OR infant* OR schoolchild* OR "school child*" OR "school*child*" OR boy* OR girl* OR teen* OR adolescen* OR Pediatric* OR Paediatric* OR Peadiatric*)) OR (AB ("Child health" OR child OR children OR child’s OR "young people" OR "young person*" OR youth* OR infant* OR schoolchild* OR "school child*" OR "school*child*" OR boy* OR girl* OR teen* OR adolescen* OR Pediatric* OR Paediatric* OR Peadiatric*)) |
| --- | --- |
| S2 | DE("Schools") OR DE("Nursery schools") OR DE("Primary education") OR DE("Secondary education") OR DE("Preschool education") OR DE("High schools") OR DE("Kindergarten") OR (TI (school* OR preschool* OR "pre school*" OR pre-school* OR nursery OR nurseries OR kindergarten* OR kindergarden*)) OR (AB (school* OR preschool* OR "pre school*" OR pre-school* OR nursery OR nurseries OR kindergarten* OR kindergarden*)) OR (TI ( "day care" OR daycare ) AND TI ( child* OR infant* )) OR (AB ( "day care" OR daycare ) AND AB ( "day care" OR daycare )) |
| S3 | (TI (close OR closed OR closure OR closures OR closing)) OR (AB (close OR closed OR closure OR closures OR closing)) |
| S4 | S2 AND S3 |
| S5 | (TI ("education access" OR "access to education")) OR (AB ("education access" OR "access to education")) OR (TI ("primary education" OR "secondary education") AND ("no access" OR "restricted access" OR "restrict*")) OR (AB ("primary education" OR "secondary education") AND ("no access" OR "restricted access" OR "restrict*")) |
| S6 | (TI ( quarantine* OR self-isolat* OR "lock down*" OR lockdown* OR "lock-down*" OR "class dismissal" OR "school dismissal*" OR "social distanc*" OR "stay at home" OR "shut down*" OR "shut-down* " OR "shutdown*" OR "staying at home")) OR (AB (quarantine* OR self-isolat* OR "lock down*" OR lockdown* OR "lock-down*" OR "class dismissal" OR "school dismissal*" OR "social distanc*" OR "stay at home" OR "shut down*" OR "shut-down* " OR "shutdown*" OR "staying at home")) |
| S7 | S4 OR S5 OR S6 |
| S8 | S1 AND S7 |
| S9 | health or well-being or wellbeing OR "Well Being" |
| S10 | S8 AND S9 |

#### eTable 2. Included outcomes

We included the following outcomes: physical health outcomes (e.g. any emergency department (ED) admission, ED admission for trauma/domestic accidents, ED admission for upper respiratory tract infection, hospital admission, management of pre-existing disease, routine vaccination, obesity; mental health outcomes (e.g. anxiety, depression, psychological distress, sense of loneliness/isolation, suicide, psychiatric admission); wellbeing outcomes (e.g sleep quality, wellbeing as perceived by children or their parents, child maltreatment); health related behaviours (e.g. physical activity, screen time, digital media use, eating behaviours, use of tobacco, alcohol, drugs).

In identified articles we also included data on: academic achievement (e.g. perceived difficulties by children/adolescents, long term outcomes such as class repetition, graduation from high school , attending higher education); and parent outcomes (e.g. lost pay/income, lost/reduction work, missed meals by children, difficulties in child care arrangement, parent anxiety, depression, distress).

Note that we did not include studies which reported only psychological mediator variables or output from structural equation models across psychological variables.

#### eTable 3. Methodological quality of cohort studies (New Castle Ottawa scale)

| **Study ID** | **Selection** | | | | **Comparability** | **Outcome** | | | **Total score** | **Overall quality** |
| --- | --- | --- | --- | --- | --- | --- | --- | --- | --- | --- |
|  | *Representative-ness of the exposed cohort* | *Selection of the non-exposed cohort* | *Ascertainment of exposure* | *Demonstration that outcome of interest was not present at start of study* |  | *Assessment of outcome* | *Follow-up long enough for outcomes to occur* | *Adequacy of follow up of cohorts* |  |  |
| Bhopal 2020 | truly representative* | drawn from the same community * | secure record* | yes * | study controls for seasonal variation* | record linkage* | yes* | all subjects accounted for* | 8/8 | high |
| Bressan 2020 | truly representative* | drawn from the same community * | secure record* | yes * | study controls for seasonal variation* | record linkage* | yes* | all subjects accounted for* | 8/8 | high |
| Chaiyachati 2020 | truly representative* | drawn from the same community * | secure record* | yes * | study controls for seasonal variation* | record linkage* | yes* | all subjects accounted for* | 8/8 | high |
| Chandir 2020 | truly representative* | drawn from the same community * | secure record* | yes * | seasonal variation not expected * | record linkage* | yes* | all subjects accounted for* | 8/8 | high |
| Chen 2020 | truly representative* | drawn from a different source | secure record* | no description | no control for confounding | self report | yes* | subjects lost to follow up unlikely to introduce bias:1.2% * | 4/8 | low |
| Ciofi Degli Atti 2020 | truly representative* | drawn from the same community * | secure record* | yes * | no: calendar period not considered | record linkage* | yes* | all subjects accounted for* | 7/8 | high |
| Cozzi 2020 | truly representative* | drawn from the same community * | secure record* | yes * | study controls for seasonal variation* | record linkage* | yes* | all subjects accounted for* | 8/8 | high |
| Dayal 2020 | truly representative* | drawn from the same community * | secure record* | yes * | study controls for seasonal variation* | record linkage* | yes* | all subjects accounted for* | 8/8 | high |
| Dyson 2020 | truly representative* | drawn from the same community * | secure record* | yes * | study controls for seasonal variation* | record linkage* | yes* | all subjects accounted for* | 8/8 | high |
| Garstand 2020 | truly representative* | drawn from the same community* | secure record* | yes * | study controls for seasonal variation* | record linkage* | yes* | all subjects accounted for* | 8/8 | high |
| Garude 2020 | truly representative* | drawn from the same community * | secure record* | yes * | study controls for seasonal variation* | record linkage* | yes* | all subjects accounted for* | 8/8 | high |
| Lazzerini 2020 | truly representative* | drawn from the same community * | secure record* | yes * | study controls for seasonal variation* | record linkage* | no | all subjects accounted for* | 7/8 | high |
| Krivec 2020 | truly representative* | drawn from the same community * | secure record* | yes * | study controls for seasonal variation* | record linkage* | yes* | all subjects accounted for* | 8/8 | high |
| Kuitunen 2020 | truly representative* | drawn from the same community * | secure record* | yes * | no: calendar period not considered | record linkage* | yes* | all subjects accounted for* | 7/8 | high |
| Iozzi 2020 | truly representative* | drawn from the same community * | secure record* | yes * | study controls for seasonal variation* | record linkage* | yes* | all subjects accounted for* | 8/8 | high |
| Isumi 2020 | truly representative* | drawn from the same community * | secure record* | yes * | study controls for seasonal variation* | record linkage* | yes* | all subjects accounted for* | 8/8 | high |
| Mann 2020 | truly representative* | drawn from the same community * | secure record* | yes * | study controls for seasonal variation* | record linkage* | yes* | all subjects accounted for* | 8/8 | high |
| Manzoni 2020 | truly representative* | drawn from the same community * | secure record* | yes * | study controls for seasonal variation* | record linkage* | yes* | all subjects accounted for* | 8/8 | high |
| McDonald 2020 | truly representative* | drawn from the same community * | secure record* | yes * | study controls for seasonal variation* | record linkage* | yes* | all subjects accounted for* | 8/8 | high |
| Odd 2020 | truly representative* | drawn from the same community * | secure record* | yes * | study controls for seasonal variation* | record linkage* | yes* | all subjects accounted for* | 8/8 | high |
| Ougrin 2020 | truly representative* | drawn from the same community * | secure record* | yes * | study controls for seasonal variation* | record linkage* | yes* | all subjects accounted for* | 8/8 | high |
| Rose 2020 | truly representative* | drawn from the same community * | secure record* | yes * | study controls for seasonal variation* | record linkage* | yes* | all subjects accounted for* | 8/8 | high |
| Sheridan 2020 | truly representative* | drawn from the same community * | secure record* | yes * | study controls for seasonal variation* | record linkage* | yes* | all subjects accounted for* | 8/8 | high |
| Sidpra 2020 | truly representative* | drawn from the same community * | secure record* | yes * | study controls for seasonal variation* | record linkage* | yes* | all subjects accounted for* | 8/8 | high |

Comparability of cohorts on the basis of the design or analysis: yes if the lockdown period has been compared with the same period (months) in the previous year(s) or if the outcomes is unlikely to be influenced by seasonal variation. No: if the lockdown period has been compared with the months immediately before lockdown and a period effect is likely to influence the outcome (seasonal variation)

Follow-up long enough for outcomes to occur**:** yes: at least 1 month FU since lockdown (lockdown lasted about two months in the majority of the countries); no: less than 1 month

##### eTable 4 Methodological quality of cross-sectional studies (Adapted New Castle Ottawa scale)

| **REFERENCE (study and year of publication)** | **Selection** | | | | **Comparability** | **Outcome** | | **Total score** | **Overall quality** |
| --- | --- | --- | --- | --- | --- | --- | --- | --- | --- |
|  | **Representativeness of the sample** | **sample size** | **Non-respondents:** | **Ascertainment of the exposure** |  | **Assessment of the outcome** | **Statistical test:** |  |  |
| CDC 2010 | truly representative* | justified and satisfactory* | unsatisfactory response rate | validated measurement* | control for important confounders* | self-report* | appropriate* | 6/7 | high |
| Children’s Society 2020 | somewhat representative° | not justified | No description | validated measurement* | no control for confounding | self-report * | not appropriate | 3/7 | low |
| Darlington 2020 | somewhat representative° | justified and satisfactory* | satisfactory response rate* | validated measurement* | control for important confounders* | self-report * | appropriate* | 7/7 | high |
| Di Giorgio 2020 | self-selected | not justified | satisfactory response rate* | validated measurement* | control for important confounders* | self-report* | appropriate* | 5/7 | medium |
| Duan 2020 | somewhat representative° | not justified | No description | validated measurement* | control for important confounders* | self-report * | appropriate* | 5/7 | medium |
| Dunton 2020 | somewhat representative° | not justified | unsatisfactory response rate | validated measurement* | control for important confounders* | self-report* | appropriate* | 5/7 | medium |
| Effler 2010 | truly representative* | justified and satisfactory* | unsatisfactory response rate | validated measurement* | control for important confounders* | self-report* | appropriate* | 6/7 | high |
| Ellis 2020 | self-selected | not justified | No description | validated measurement* | control for important confounders* | self-report * | appropriate* | 4/7 | medium |
| Falkingham 2020 | truly representative* | justified and satisfactory* | No description | validated measurement* | control for important confounders* | self-report* | appropriate* | 6/7 | high |
| Garcia de Avila 2020 | self-selected | not justified | No description | validated measurement* | control for important confounders* | self-report * | appropriate* | 4/7 | medium |
| Gelardi 2020 | somewhat representative° | justified and satisfactory* | satisfactory response rate* | validated measurement* | no control for confounding | self-report* | appropriate* | 6/7 | high |
| Gift 2010 | truly representative* | justified and satisfactory* | unsatisfactory response rate | validated measurement* | no control for confounding | self-report * | appropriate* | 5/7 | medium |
| Ki 2020 | somewhat representative° | not justified | No description | validated measurement* | control for important confounders* | self-report * | appropriate* | 5/7 | medium |
| Kilincel 2020 | self-selected | not justified | No description | validated measurement* | control for important confounders* | self-report* | appropriate* | 4/7 | medium |
| Jeasmin 2020 | selected group | justified and satsfactory* | satisfactory response rate* | validated measurement* | control for important confounders* | self-report * | appropriate* | 6/7 | high |
| Johnson 2008 | truly representative* | justified and satisfactory* | satisfactory response rate* | validated measurement* | no control for confounding | self-report * | appropriate* | 6/7 | high |
| Levita 2020 | truly representative* | justified and satisfactory* | satisfactory response rate* | validated measurement* | no control for confounding | self-report * | not appropriate | 5/7 | medium |
| Lopez Bueno 2020 | somewhat representative° | justified and satisfactory* | satisfactory response rate* | validated measurement* | control for important confounders* | self-report* | appropriate* | 7/7 | high |
| Lynn 2020 | truly representative* | justified and satisfactory* | unsatisfactory response rate | validated measurement* | not applicable | self-report * | appropriate* | 6/6 | high |
| Marino 2020 | self-selected | not justified | No description | validated measurement* | no control for confounding | self-report* | appropriate* | 3/7 | low |
| Roland 2020 | truly representative* | justified and satisfactory* | satisfactory response rate* | validated measurement* | not applicable | self-report * | appropriate* | 6/6 | high |
| Roy 2020 | somewhat representative° | justified and satisfactory* | satisfactory response rate* | validated measurement* | no control for confounding | self-report* | appropriate* | 6/7 | high |
| Russel 2020 | selected group | not justified | No description | validated measurement* | control for important confounders* | self-report* | appropriate* | 4/7 | medium |
| Timperio 2009 | truly representative* | not justified | unsatisfactory response rate | validated measurement* | control for important confounders* | self-report* | appropriate* | 5/7 | medium |
| Tsai 2017 | truly representative* | justified and satisfactory* | unsatisfactory response rate | validated measurement* | control for important confounders* | self-report* | appropriate* | 6/7 | high |
| Xie 2020 | selected group | not justified | satisfactory response rate* | validated measurement* | no control for confounding | self-report * | appropriate* | 4/7 | medium |
| Watson 2020 (A-D)) | self-selected | not justified | No description | validated measurement* | no control for confounding | self-report * | not appropriate | 2/7 | low |
| Zheng 2020 | selected group | not justified | satisfactory response rate* | validated measurement* | control for important confounders* | self-report* | appropriate* | 5/7 | medium |
| Zheteyeva, Y. 2017 | truly representative* | justified and satisfactory* | unsatisfactory response rate | validated measurement* | control for important confounders* | self-report* | appropriate* | 6/7 | high |
| Zhou 2020 a | somewhat representative° | justified and satisfactory* | satisfactory response rate* | validated measurement* | control for important confounders* | self-report * | appropriate* | 7/7 | high |
| Zhou 2020 b | somewhat representative° | justified and satsfactory* | satisfactory response rate* | validated measurement* | control for important confounders* | self-report * | appropriate* | 7/7 | high |
| Zhou 2020 c | somewhat representative° | justified and satsfactory* | satisfactory response rate* | validated measurement* | control for important confounders* | self-report * | appropriate* | 7/7 | high |

| **WAS FROM CASP** | **Aim clearly stated** | **Qualitative methodology appropriate** | **Research design appropriate to address the aim of the research** | **Recruitment strategy appropriate** | **data collected in a way that addressed the research issue** | **Relationship between researcher and participants adequately considered** | **Ethical issues taken into consideration** | **Data analysis sufficiently rigorous** | **Clear statement of findings** | **Total score** | **Overall quality** |
| --- | --- | --- | --- | --- | --- | --- | --- | --- | --- | --- | --- |
| Segre 2020 | yes | yes | yes | yes | yes | no | yes | yes | yes | 8/9 | high |

Representativeness of the sample: truly representative: all or random sample of the target population; somewhat representative: non random sampling; selected group: subgroup of target population selected by researchers; self-selected group: participants recruited though online advertising

Sample size justified and satisfactory: if the study use data from a large national survey or if the study provides a sample size calculation. Not justified in the other cases

Non responders: satisfactory if respondents are at least 70% or if comparability is described

Statistical analysis appropriate: for descriptive studies numerator and denominator clearly reported, and percentages given with confidence intervals; for studies that evaluate association statistical analysis described in the methods section.

##### eTable 5. Methodological quality of uncontrolled pre-post studies (NHLBI checklist)

| **Study ID** | **Objective clearly stated** | **Eligibility criteria defined** | **Participants representative of the population** | **All eligible participants enrolled** | **Sample size satisfactory** | **Exposure clearly described** | **Outcomes specified and clearly described** | **Blinding outcome assessor** | **Follow up rate** | **Statistical analysis appropriate** | **Multiple outcome measures** | **Group-level interventions and individual-level outcome** | **Total** | **Overall quality** |
| --- | --- | --- | --- | --- | --- | --- | --- | --- | --- | --- | --- | --- | --- | --- |
| Angoulvant 2020 | yes | yes | yes | yes | yes | yes | yes | NA | yes | yes | yes | NA | 10/10 | high |
| Bandyopadhyay 2020 | yes | no | nr | nr | nr | yes | yes | NA | CD | no | no | NA | 3/10 | low |
| Baron 2020 | yes | yes | yes | yes | yes | yes | yes | NA | yes | yes | yes | NA | 10/10 | high |
| Baysun 2020 | yes | no | no | no | no | yes | yes | NA | CD | no | no | NA | 3/10 | low |
| Christoforidis 2020 | yes | yes | no | nr | no | yes | yes | NA | CD | yes | no | NA | 5/10 | low |
| Della Giulia 2020 | yes | no | no | nr | no | yes | yes | NA | yes | yes | no | NA | 5/10 | low |
| Gallagher 2020 | yes | yes | no | nr | yes | yes | yes | NA | no | yes | no | NA | 6/10 | medium |
| Heymann 2004,2009 | yes | yes | yes | yes | yes | yes | yes | NA | yes | yes | no | NA | 9/10 | high |
| Martinelli 2020 | no | no | no | nr | no | yes | yes | NA | yes | yes | no | NA | 4/10 | low |
| Nastro 2020 | yes | yes | no | nr | no | yes | yes | NA | yes | yes | no | NA | 6/10 | medium |
| Pearcey 2020 | yes | yes | no | nr | yes | yes | yes | NA | no | yes | mo | NA | 6/10 | medium |
| Pearcey 2020 b | yes | yes | no | nr | yes | yes | yes | NA | no | yes | mo | NA | 6/10 | medium |
| Pietrobelli 2020 | yes | yes | no | nr | no | yes | yes | NA | yes | yes | no | NA | 6/10 | medium |
| Tittel 2020 | yes | yes | yes | yes | yes | yes | yes | NA | yes | yes | yes | NA | 10/10 | high |
| Widnall 2020 | yes | no | nr | nr | yes | yes | yes | NA | no | no | no | NA | 4/10 | low |

NA: not applicable; CD: can’t tell; NR: not reported. Follow up rate: adequate if lost are <80%

#### eTable 6. Methodological quality of modelling studies (adapted from ISPOR checklist)

| **Study ID** | **Design: clear description of modelling objective and scope** | **Assumption in the model described and appropriate** | **External and internal validation performed and described** | **Face validity: the model considers all relevant aspects of population, behaviors, setting** | **Data: data used to inform model adequate** | **Analysis: adequate assessment of the effects of uncertainty** | **Total score** | **Overall quality** |
| --- | --- | --- | --- | --- | --- | --- | --- | --- |
| AN 2020 | yes | yes | no | yes | yes | yes | 5/6 | high |

##### eTable 7. Health service use 7A. ED attendances

| ***Study ID*** | ***N*** | ***Total attendances*** | | | ***fever, respiratory infection*** | | | ***domestic injuries*** | | | ***All trauma or injury*** | | |
| --- | --- | --- | --- | --- | --- | --- | --- | --- | --- | --- | --- | --- | --- |
|  |  | ***pre*** | ***lockdown*** | ***difference*** | ***pre*** | ***lockdown*** | ***difference*** | ***pre*** | ***lockdown*** | ***difference*** | ***pre*** | ***lockdown*** | ***difference*** |
| Angoulvant 2020 | 871 543 PED visits |  |  | −68.0% [−81.2 to −55.8] |  |  |  |  |  |  |  |  |  |
| Bressan 2020 | 3713 | 8 March- 20 April 2019: 2917 | 8 March -20 April 2020: 796 | -72.7% |  |  |  | 8 March- 20 April 2019: 148 | 8 March -20 April 2020: 178 | IRR 1.2 (1.0 to 1.5) p:0.09 |  |  |  |
| Chaiyachati 2020 | 29496 | March 23,-April 21, 2017-2019: total: 8849 per year; daily visits : 286(SDS42) | March 23-April 21, 2020: total: 2948; daily visits 95(SD16) | -66.8% in daily visits; p<0.001 | March 23- April 21, 2017-2019: total: 1229 (SD 233) | March 23-April 21, 2020: total: 394 | -67.9%; p<0.001 |  |  |  | March 23,-April 21, 2017-2019. total: 1274 (SD102) | March 23- April 21, 2020: total:449 | p<0.001 |
| Ciofi Degli 2020 | 18825 | 1 January-19 February. (50 days) N.visits: 11956. Daily visits: 239.1 (SD 28.4) | 20 February- 10 March(partial school closure) daily visits: 180.2 (SD:39.2); 11 March-20 April (school closure+ lockdown): daily visits 79.6 (SD 10.4) | Partial School closure: -24.6% Lockdown: -66.7% ; p<0.05 | 1 January-19 February: daily visits 82.4 (SD 23.7) | 20 February- 10 March: daily visits 55.3(SD 16.1); 11 March-20 April: daily visits 18.7 (SD 5.4) | School closure: -32.8% p<0.001 Lockdown: -77.3% p<0.001 | 1 January-19 February:daily visits 9.6 (SD 4.2) | 20 February- 10 March: daily visits 11.8(SD 3.9); 11 March-20 April:daily visits 6.5 (SD 7.1) | p<0.001 | 1 January-19 February: daily visits 41.9 (SD 9.9) | 20 February- 10 March: daily visits 38.4 (SD 7.3) 11 March-20 April:: daily visits 23.8 (SD 6.9) | p<0.001 |
| Cozzi 2020 | 3362 | 2019: 2866 prelockdown 2020: 2719 | 9 March - 13 April: 646 | From 2019: -77.5% From prelockdown 2020: -76.3% | 2019:171 (6%) pre-lockdown: 302 (11.1%) | 69 (10.7%) | From 2019: p<0.00001 From pre-lockdown: NS |  |  |  | 2019: 703 (24.5%) Pre-lockdown: 613 (22.5%) | 150 (23.5%) | NS for proportions due to injuries |
| Dyson 2020 | 146 |  |  |  |  |  |  |  |  |  | 17 | 27 | p=0.35 |
| Garude 2020 | 37 |  |  |  |  |  |  |  |  |  |  |  |  |
| Iozzi 2020 | 2956 | 2310 | 646 | -73% | fever: 73 Pneumonia: 44 | fever: 36 Pneumonia: 23 |  |  |  |  | 77 (3.3% of cases) | 124 (19.2% of cases) | +61% |
| Kuitunen 2020 |  | Hospital n 1 :total n. visits: 615, median daily visits: 19. Hospital n 2: total n. visits: 266,median daily visits: 9 | Hospital n.1: total n. vists: 211,median daily visits: 7.Hospital n 2: total n. visits: 92, median daily visits: 2.5 | Hospital 1: -63.2%, p<0.001 Hospital 2: -72.2%, p<0.001 | Hospital 1 upper respiratory 221, lower respiratory 33; Hospital 2 upper respiratory 81, lower respiratory 58. | Hospital 1 upper respiratory 71, lower respiratory 16; Hospital 2 upper respiratory 26, lower respiratory 9 | Hospital 1 : p=0.65 Hospital 2: p=0.04 |  |  |  |  |  |  |
| Manzoni 2020 | 1654 | total visits: 1428; mean daily visits: 23.4 | total visits: 226; mean daily visits: 3.4 | -84%, p<0.001 | mean daily visits: 5.5 (23.5% of all attendances) | mean daily visits: 0.4 (11.9% of attendances) | -92%, p<0.001 |  |  |  | mean daily visits: 4.6 (19.8% of total) | mean daily visits: 1.1 (29.2% of total) | -76%, p=0.001 |
| Mann 2020 | nr |  |  |  |  |  |  | Burns: 2019: 83 of total of 12 599 ED presentations (0.7%) | Burns: 2020:64 of total of 5031 (1.3%) |  |  |  |  |
| Rose 2020 PP | 3545 | 2019: 42838 total attendances: green code (lowest risk): 2251 (53.1%); yellow code: 1301 (30.7%) orange code: 575 (13.6%); red code (highest risk): 41 (0.97%) | 2020: total 453, green code: 214 (47.4%); yellow code: 136 (30.1%); orange code:89 (19.7%); red code: 5( 1.11%) | 89.3% reduction in total attendances. Proportional increase in two highest risk categories 6.3% (p<0.001), proportional decrease in bottom two categories 4.7% (p=0.07) |  |  |  |  |  |  |  |  |  |
| Sidpra 2020 | 10 in 2020 NR in the previous years |  |  |  |  |  |  |  |  |  | suspected abusive head trauma (AHT) 2017-2019: mean 0.67 cases per month | 23 March and 23 April 2020: n=10 | +1493% |
| Lazzerini 2020 | nr | 1-27 March 2019: n=9276 | 1-27 March 2020: n=2008 | -78% |  |  |  |  |  |  |  |  |  |
| ***PRE COVID*** |  | ***Before*** | ***During*** |  |  |  |  |  |  |  |  |  |  |
| Heymann 2004 & 2009 | 186094 | 3.2/1000 | 2.5/1000 | -28% (95% CI 11–46%); p<0.001 | diagnosis of URI (not in ED). 44/1000. | 24.7/1000 | p<0.001. |  |  |  |  |  |  |

Notes:

nr: not reported

PED: Paediatric emergency department

NS: not significant

Difference: - reduction; + increase : in lockdown compared with historical control periods

PICU: Paediatric intensive care unit

URI: upper respiratory infection

##### 7B. Hospital admissions

| ***Study ID*** | ***N*** | ***Total admissions*** | | | ***domestic injuries*** | | | ***Injuries or trauma*** | | | ***fever, respiratory infection*** | | |
| --- | --- | --- | --- | --- | --- | --- | --- | --- | --- | --- | --- | --- | --- |
|  |  | ***pre*** | ***lockdown*** | ***difference*** | ***pre*** | ***lockdown*** | ***difference*** | ***pre*** | ***lockdown*** | ***difference*** | ***pre*** | ***lockdown*** | ***difference*** |
| Angoulvant 2020 | 871 543 PED visits |  |  | −45.0% [−57.0 to −32.4] |  |  |  |  |  |  |  |  |  |
| Bressan 2020 | 3713 |  |  |  | 8 March- 20 April 2019:4 | 8 March -20 April 2020: 20 | IRR 5.0 (1.7 to 14.6) p: 0.003 |  |  |  |  |  |  |
| Ciofi Degli 2020 | 18825 | 20 February-20 April: N. urgent admission:1530; daily urgent admissions 30.6 (SD 6.8) | February- 10 March: daily admission: 27.7 (SD 6.0). 11 March-20 April):daily admissions 21.2 (SD 4.9) | School closure: -9.5% Lockdown: -30.7% | 1 January-19 February: daily 0.7 (SD 1) | 20 February- 10 March: daily 1 (SD 1.1) ; 11 March-20 April : daily 0.7 (SD 1.1) | NS | 1 January-19 February: daily 3.1 (SD 2.0) | 20 February- 10 March: daily 3.4 (SD 2); 11 March-20 April: daily 3 (SD 1.8) | NS | 1 January-19 February: daily 8.8 (SD 3.7) | 20 February- 10 March: daily 6.8 (SD 2.6); 11 March-20 April : daily 5.1 (SD 2.7) | School closure: -22.7% p<0.001 Lockdown: -42.0% p<0.001 |
| Cozzi 2020 | 3362 | 2019:80 Prelockdown 2020: 62 | 48 | From 2019: -40.0-%, p <0.0001  From prelockdown: 22.6%, p <0.0001 |  |  |  | 2019: 20 Prelockdown: 25 | 17 | From 2019: 5%, p<0.0001 From prelockdown: 32%, p<0.0001 |  |  |  |
| Dyson 2020 | 146 | neurosurgical: 68 | neurosurgical: 78 |  |  |  |  |  |  |  |  |  |  |
| Garude 2020 | 37 |  |  |  |  |  |  | hand trauma: 31 | hand trauma: 6 | -80.6% |  |  |  |
| Krivec 2020 | nr |  |  |  |  |  |  |  |  |  | 2017-2019: mean: 51 | 20 | -60.8% |
| Kuitunen 2020 |  | Hospital 1 : 29 (4.7% of ED attendances). Hospital 2: : 100 (38% of attendances) | Hospital 1: 16 (7.6% of attendances) Hospital 2: 40 (44% of attendances) | Hospital 1: -44.8%, p=0.13  Hospital 2: -60.0%, p=0.32 |  |  |  |  |  |  |  |  |  |
| Manzoni 2020 | 1654 | mean daily admission:1.2 (5% of ED presentations) | mean daily admission:0.3 (8.1% of ED admissions) | -75%; p=0.02 |  |  |  |  |  |  |  |  |  |
| Rose 2020 PP | 3545 | 2019: ward 713 (16.8% of ED attendances); ICU:4 (0.09%) | 2020: ward 102 (22.6% of attendances); ICU: 1 (0.22%) | -85.7% for total admissions. Increase in proportion of ED presentations admitted of 13.2% (CI 9.11 - 17.7%, p<0.001). |  |  |  |  |  |  |  |  |  |
| ***PRE COVID*** |  |  |  |  |  |  |  |  |  |  |  |  |  |
| Heymann 2004/ 2009 | 186094 | 1.6/1000 | 1.7/1000 |  |  |  |  |  |  |  |  |  |  |

##### 7C. Hospitalisations for specific conditions

| ***Study ID*** | ***N*** | ***Asthma*** | | | ***Type 1 diabetes*** | | | ***New presentations of type 1 diabetes*** | | | ***Delayed presentations*** |
| --- | --- | --- | --- | --- | --- | --- | --- | --- | --- | --- | --- |
|  |  | ***pre*** | ***lockdown*** | ***difference*** | ***pre*** | ***lockdown*** | ***difference*** | ***pre*** | ***lockdown*** | ***difference*** |  |
| Dayal 2020 | nr |  |  |  | April 2019 - March 2020: mean 19 (range 12-28) per month | April 2020: 4 | -79% | April 2019 - March 2020:mean 12 (range 7-19) 15% with severe ketoacidosis | April 2020: 3, all with severe ketoacidosis | -75% | All cases with severe ketoacidosis were delayed presentations |
| Krivec 2020 | nr | 2017-2019: mean: 8.3 | 2 | -75.9% |  |  |  |  |  |  |  |
| Tittel 2020 | 4628 |  |  |  |  |  |  | type 1 diabetes incidence increasedfrom16.4[14.7–18.2] in 2011 to 22.2 [20.3–24.2] in 2019 | 2020:incidence 23.4 [21.5–25.5] | Incidence in 2020 did not differ significantly from the trend predicted (22.1 [20.4–23.9]) |  |
| Lazzerini 2020 | nr |  |  |  |  |  |  |  |  |  | 12 serious cases of delayed access to healthcare (predominantly sepsis; ketoacidosis; cancer presentations), of whom 6 required intensive care and 4 died |
| Roland 2020 | 1349 |  |  |  |  |  |  |  |  |  | 3.8% (n=51) felt to have some delay in presentation (3.0% due to parental issues, 0.8% due to professional advice); 6/51 (11.8%) were admitted to hospital, with 1 of these admitted to PICU. Uncertain re delay in 2.7% (n=36) |
| Lynn 2020 | 2433 paediatricians |  |  |  |  |  |  |  |  |  | 32% of those working in urgent care and 18% of those working on hospital wards had witnessed delayed presentations during the first month of lockdown. 9 deaths were considered to be related to delayed presentation |

##### eTable 8. Vaccination

| **Study id** | **exposure** | **n** | **Pre** | **During lockdown** | **Difference** |  |
| --- | --- | --- | --- | --- | --- | --- |
| Chandir 2020 | L | 71324 | Sept 23, 2019–March 22, 2020: 5184 | March 23–May 9, 2020: 2450 | 2734 children per day missed routine immunisation (-52.8%) | This decline can be attributed to a combination of demand and supply factors. Restrictions on movement and concerns around COVID-19 transmission might have prevented caregivers from accessing immunisation services. Of all the 321 operational immunisation centres, 50 (16%) had no client flow. The supply side was also adversely affected, as 48 (18%) of all 271 immunisation centres were closed. Our analysis depicts the widening schism in the proportion of immunisation coverage between the different wealth quintiles as the more affluent neighbourhoods of the city have been less affected by the restrictions. Previous surveys have shown existing inequities in the proportion of full immunisation coverage between the lowest and the highest wealth quintiles in the country, which are now being exacerbated |
| McDonald 2020 PP | L | 136698 hexavalent 1 vaccine; 127173 MMR1 vaccine | hexavalent 1: 69,568 MMR1:65,341 | hexavalent 1: 67,116 MMR1:61,832 | hexavalent 1: -3.5 (-3.7 to -3.4) MMR1: -3.7 (-3.8 to -3.6) | Decreasing birth rates may plausibly explain the gradually falling hexavalent vaccination counts,(7) and migration could play a role too, but these cannot explain the size and timing of the changes in MMR vaccination |

Notes

SC+L: school closure + lockdown

L: lockdown

##### eTable 9. Mental health. 9A Overall distress, anxiety and depression

| **Study** | **Exposure** | **N** | **Psychological distress / difficulties** | | | **Anxiety** | | | **Depression** | | |
| --- | --- | --- | --- | --- | --- | --- | --- | --- | --- | --- | --- |
|  |  |  | **pre** | **during lockdown** | **difference** | **pre** | **during lockdown** | **difference** | **pre** | **during lockdown** | **difference** |
| Russel 2020 | SC+L | 420 |  | National Institutes of Health Toolkit’s emotion resources: mean score 23.8 (SD 6.9) which is >50th percentile (indicates moderate child stress). Higher child reported stress in parents with financial problems. |  |  |  |  |  |  |  |
| Sheridan 2020 | SC+L | nr |  |  |  |  |  |  |  |  |  |
| Chen 2020 | SC+L | 7772 |  |  |  |  | Generalized Anxiety Disorder-7 (GAD-7 ): Wuhan:821/2850 (28.8%); other cities 1270/4922 (25.80%) | p = .004 |  | Patient Health Questionnaire-9 (PHQ-9): Wuhan:1245/2850 (43.7%); other cities 2089/4922 (42.4%) | p = .286 |
| Di Giorgio 2020 | SC+L | 245 |  |  | Increases in SDQ subscales: emotional problems p=0.011, conduct problems p=0.04, hyperactivity/inattention p<0.0001 |  |  |  |  |  |  |
| Kilincel 2020 | SC+L | 745 |  |  |  |  | State Trait Anxiety Inventory (STAI): STAI-S: mean 43.17 ± 5.86. STAI-T: mean: 51.53 ± 5.19 |  |  |  |  |
| Roy 2020 PP | SC+L | 1065 |  |  | Reported stress: increase drastically: 11.7%, incerased mildly:40.2%, decreased mildly:35.2%, derceased drastically: 12.9% |  |  |  |  |  |  |
| Segre 2020 PP | SC+L | 82 |  |  |  |  | Anxiety scale of Trauma and Symptom checklist for children” (positive for anxiety if the total score was.)mean score was 11.56 (2.65 s.d.); 78 % were high scorer (scored ≥ 10) |  |  | Mood scale of 6 questions: Mean score: 3.43 (SD1.6). high scorers (≥ 4)=43.9% |  |
| Zheng 2020 PP | SC+L | 1620 |  |  |  |  | Social Anxiety Scale for Children (SASC). Mean 3.90 ± 3.73, significantly higher than the norm scores. High scorers for social anxiety= 279 (17.2%) |  |  | Depression Self-rating Scale for Children (DSRSC). Mean: 5.67 ± 4.97 lower than norm score. High scorers for depressive symptoms = 102 (6.3%) |  |
| Children’s Society 2020 | SC+L | 150 |  |  |  |  |  |  |  |  |  |
| Xie 2020 | SC+L | 1784 |  |  |  |  | anxiety symptoms measured by the Screen for Child Anxiety Related Emotional Disorders: 337 (18.9%) |  |  | depressive symptoms measured by the Children’s Depression Inventory–Short Form (CDI-S): 403 (22.6%) |  |
| Widnal 2020 | SC+L | n=1047longitudinal n= 721-770 |  |  |  | Hospital Anxiety and Depression Scale (HADS) ; high scorer: (>=9): Girls: 54%; boys: 26% | High scorer: girls 45%; boys 18% |  | Hospital Anxiety and Depression Scale (HADS).High scorer (>=7): Girls 31% Boys 21% | High scorer: girls 34%, boys 19% |  |
| Odd 2020 | SC+L | 51 |  |  |  |  |  |  |  |  |  |
| Watson 2020 | SC+L | 11228 |  | SDQ: 41% of 4-7 year olds had borderline or high scores for hyperactivity / inattention, 43% for conduct problems and 37% for emotional problems, higher than the 20% expected in community samples |  |  |  |  |  | mood worse than before: 47% |  |
| Levita 2020 | SC+L | 1001 |  |  |  |  | Hospital Anxiety and Depression Scale (HADS): anxiety in 53.3% of girls and 44% of boys aged 13-18. Revised Child Impact Events Scale (CRIES-8): high score >17: 53.3% of girls and 44% of boys |  |  | HADS: depressive symptoms 19.4% of girls and 21.9% of boys |  |
| Gallagher 2020 | SC+L | 194 parents, 58 adolescents |  |  | SDQ: no significant change in mean scores for emotional, behavioural or restlessness/inattention difficulties amongst 4-11 year olds (parent report) nor in parental or adolescent report of emotional or behavioural difficulties amongst 12-18 year olds. Parents/carers of children with a pre-existing mental health or neurodevelopmental conditions reported a statistically significant reduction in their child’s emotional difficulties over a 1-month period as COVID-19 restrictions progressed |  |  |  |  |  |  |
| Pearcey 2020 | SC+L | 972 |  |  | SDQ: no changes in emotional difficulties but reduction in restlessness/inattention difficulties over 1 month in lockdown, whilst behavioural difficulties reduced significantly in boys but not girls during lockdown |  |  |  |  |  |  |
| Nastro 2020 | SC+L | 71 |  |  |  |  | Promis anxiety questionnaires: 73% did not show anxiety | small non-significant decreases in somatic symptoms and anxiety during lockdown compared with a year previously |  |  |  |
| Pearcey 2020 B | SC+L | 2890 parents and 572 adolescents |  |  | SDQ: Parents reported emotional, behavioural and restlessness/inattention difficulties increased significantly during lockdown in 4-10 year olds. In 11-16yo, parents’ reported that emotional difficulties decreased and restlessness/inattention difficulties increased significantly although young people themselves reported no change in difficulties |  |  |  |  |  |  |
| Eillis 2020 | SC+L | 1054 |  |  |  |  |  |  |  | Brief Symptom Inventory: 28% reported symptoms across all 6 items |  |
| Garcia 2020 | SC+L | 289 |  |  |  |  | Children’s Anxiety Questionnaire (CAQ) high score (>9)=19.4%. Numerical Rating Scale (NRS) high score (>7) = 21.8%. No differences between sexes. Higher anxiety in those with parent/carer with only elementary school education (p=0.02) |  |  |  |  |
| Yeasmin 2020 | SC+L | 384 |  | Cluster analysis from Revised Child Anxiety and Depression Scale (RCADS), Generalized Anxiety Disorder scale (GAD),and Child Behavior Checklist (CBCL): 43% of child had subthreshold mental health disturbances, 30.5% had mild disturbances, 19.3% suffered from moderate disturbance ; 7.2% suffered from severe disturbance |  |  |  |  |  |  |  |
| Zhou 2020 | SC+L | 4805 |  |  |  |  |  |  |  | Center for Epidemiologic Studies Depression Scale (CES-D): high score (>15) 39.5% |  |
| Duan 2020 | SC+L | 3613 |  |  |  |  |  |  |  | Child Depression Inventory (CDI): high scorer (>=19)=22.3% |  |
| Qi 2020 | SC+L | 9554 |  |  |  |  | Generalized anxiety disorder (GAD-7) scale: high scorer: 19.0% ((18.2, 19.8) |  |  |  |  |
| Zhou 2020 | SC+L | 8079 |  |  |  |  | Generalized Anxiety Disorder scale (GAD-7): moderate-severe symptoms 10.4% |  |  | Patient Health Questionnaire (PHQ-9): moderate-severe symptoms 17.3% |  |

Notes:

SDQ: Strengths and Difficulties Questionnaire

SC+L: school closure + lockdown

L: lockdown

##### 9B. Psychiatric health service use, self-harm and suicide, social support and executive function

| **Reference** | **Exposure** | **N** | **Social support and connection** | **Suicide and self-harm** | | | **Psychiatric admissions or presentations** | | | **Executive function** | | |
| --- | --- | --- | --- | --- | --- | --- | --- | --- | --- | --- | --- | --- |
|  |  |  |  | **pre** | **during lockdown** | **difference** | **pre** | **during lockdown** | **difference** | **pre** | **during lockdown** | **difference** |
| Isumi 2020 | SC+L | nr |  |  |  | no significant change of suicide rate during March-May from 2018 to 2020: incidence rate ratio (IRR) = 1.15 ( 0.81,1.64) |  |  |  |  |  |  |
| Ougin 2020 | SC+L | 3141 |  |  |  |  | Inpatient admissions: Mean 2017-2019: March=385.7. April =385 | March 2020: 217 April 2020: 244 | -40.2% |  |  |  |
| Russel 2020 | SC+L | 420 |  |  |  |  |  |  |  |  |  |  |
| Sheridan 2020 | SC+L | nr |  | Self-harm presentations: 46 | 16 | -65.2% |  |  | ED mental health presentations declined to just under half expected number for March and April |  |  |  |
| Chen 2020 | SC+L | 7772 |  |  |  |  |  |  |  |  |  |  |
| Di Giorgio 2020 | SC+L | 245 |  |  |  |  |  |  |  | Behavior Rating Inventory of Executive Functions— preschool (BRIEF-P): self-control difficulties 14.3% | 21.20% | +48.3%; p<0.001 |
| Kilincel 2020 | SC+L | 745 | UCLA loneliness scale: mean: 41.89 ± 9.81 |  |  |  |  |  |  |  |  |  |
| Roy 2020 PP | SC+L | 1065 |  |  |  |  |  |  |  |  |  |  |
| Segre 2020 PP | SC+L | 82 |  |  |  |  |  |  |  |  |  |  |
| Zheng 2020 PP | SC+L | 1620 |  |  |  |  |  |  |  |  |  |  |
| Children’s Society 2020 | SC+L | 150 | Likert measure of coping during lockdown (0 =not coped very well to 10=coped very well). Overall, 84% scored above midpoint i.e. copied well. For social ìsolation: 17% below midpoint; For not being able to see friends: 37% below midpoint |  |  |  |  |  |  |  |  |  |
| Xie 2020 | SC+L | 1784 |  |  |  |  |  |  |  |  |  |  |
| Widnal 2020 | SC+L | n=1047, longitudinal n= 721-770 | School connectedness scores increased; girls: pre 3.0 lockdown 3.3; boys pre 3.2 lockdown 3.5. Those with low pre-pandemic school connection showed greater reduction in anxiety scores but little change in depression scores. Peer and family connection scores changed little pre-pandemic to lockdown. Anxiety and depression scores increased most in those with low family and low peer scores pre-pandemic. |  |  |  |  |  |  |  |  |  |
| Odd 2020 | SC+L | 51 |  | 1st January to 22nd March 2020 - 82 days) n=26. 1st April 2019 and 17th May 2019 n=14 | 23rd March 2020-17th May 2020 (56 days) n=25. 1st April 2020-17th May 2020 n=21 | Lockdown vs pre lockdown: Rate ratio (RR) 1·41 (95% CI 0·80-2·46) p=0·230. 2020 vs 2019: RR 1·50 (95% CI 0·75-2·99); p=0· 249 |  |  |  |  |  |  |
| Watson 2020 | SC+L | 11228 |  |  |  |  |  |  |  |  |  |  |
| Levita 2020 | SC+L | 1001 |  |  |  |  |  |  |  |  |  |  |
| Gallagher 2020 | SC+L | 194 parent, 58 adolescents |  |  |  |  |  |  |  |  |  |  |
| Pearcey 2020 | SC+L | 972 |  |  |  |  |  |  |  |  |  |  |
| Nastro 2020 | SC+L | 71 |  |  |  |  |  |  |  |  |  |  |
| Pearcey 2020 B | SC+L | 2890 parents and 572 adolescents |  |  |  |  |  |  |  |  |  |  |
| Eillis 2020 | SC+L | 1054 |  |  | moderate or severe suicidal ideation in 17.5%; compared with approximately 6% in normative data |  |  |  |  |  |  |  |
| Garcia 2020 | SC+L | 289 |  |  |  |  |  |  |  |  |  |  |
| Yeasmin 2020 | SC+L | 384 |  |  |  |  |  |  |  |  |  |  |
| Zhou 2020 | SC+L | 4805 |  |  |  |  |  |  |  |  |  |  |
| Duan 2020 | SC+L | 3613 |  |  |  |  |  |  |  |  |  |  |
| Qi 2020 | SC+L | 9554 |  |  |  |  |  |  |  |  |  |  |
| Zhou 2020 | SC+L | 8079 |  |  |  |  |  |  |  |  |  |  |

SC+L: school closure + lockdown

L: lockdown

##### 9C. Wellbeing

| **Study** | **Exposure** | **N** | **Pre** | **During lockdown** | **difference** |
| --- | --- | --- | --- | --- | --- |
| Children’s Society 2020 | SC+L | over 2000 household |  | Life satisfaction: 18% below midpoint. Warwick Edinburgh Mental Well-being Scale (WEMWBS) (only 13-17 years ): Low score. (<43):26.9% |  |
| Widnal 2020 | SC+L | n= 721-770 | Warwick-Edinburgh Wellbeing Scale (WEMWBS); Note the UK average score on this questionnaire for 13-16 year olds is 48.8. October 2019: girls:mean 44; boys: mean 48.7 | Girls: mean: 45; Boys mean: 51 | Not tested |

SC+L: school closure + lockdown

L: lockdown

##### eTable 10. Child abuse and protection

| **Study** | **Exposure** | **N** | **Child maltreatment allegations** | | | **Medical examination as part of a child protection assessment** | | |
| --- | --- | --- | --- | --- | --- | --- | --- | --- |
|  |  |  | **Pre** | **During lockdown** | **Difference** | **Pre** | **During lockdown** | **Difference** |
| Bhopal 2020 | SC+L | 407 |  |  |  | March 2018: 36 , 2019: 43. January-April 2018: 152, January-April 2019: 156 | March 2020: 28. January-April: 99 | March comparison: -29.1% Jan-April comparison: 35.7% |
| Garstand 2020 PP | SC+L | 191 | Referrals: 2018=78, 2019=75 School as source of referral:2018=47.4%, 2019=52.1% | Referrals: 2020=47. School as source of referral 2020: 25.5% | Reduction in referrals of 39% (95% CI 14%-57%). Schools as source of referral: p=0.015 |  |  |  |
| Baron 2020 | SC+L | 13,132 county-by-month observations |  |  | An econometric model showed the predicted number of allegations was similar to the actual number for every available month in the 2019–20 academic year, except for March and April 2020, with reductions of 4200 and 10,700 allegations respectively relative to the counterfactual. Overall this is -27%. They use a detailed dataset of school district staffing and spending to show that the observed decline in allegations was largely driven by school closures. |  |  |  |

SC+L: school closure + lockdown

L: lockdown

##### eTable 11. Sleep

| **Study** | **Exposure** | **N** | **Duration (hours/ day)** | | | **Quality** | | |
| --- | --- | --- | --- | --- | --- | --- | --- | --- |
|  |  |  | **Pre** | **During lockdown** | **Difference** | **Pre** | **During lockdown** | **Difference** |
| Pietrobelli 2020 | SC+L | 41 | 8.46 ± 0.85 | 9.11 ± 1.10 | 0.65 ± 1.29, p=0.003 |  |  |  |
| Della Giulia 2020 | SC+L | 37 |  |  |  |  |  | Growth models showed that early in lockdown parents reported more challenging bedtime routines, and children’s sleep quality and duration decreased. This was followed by a stabilization of routine and the quantity and quality of sleep also stabilised although at a poorer level than initially |
| Di Giorgio 2020 | SC+L | 245 |  |  |  | Sleep Disturbance Scale for Children (SDSC): sleep difficulties (> 39)= 41.6% | % of chidern with sleep difficulties: 44.7% | P. 0.4 OR: 1.23 (0.743-2.04) |
| Falkingham 2020 | SC+L | 895 |  | Responses: ‘much more than usual’ to lose sleep: 5.2%; sleep loss during the coronavirus pandemic: 29.2%; % new occurrence of sleep loss during lockdown: 25% |  |  |  |  |
| Lopez Bueno 2020 | SC+L | 860 | before confinement: 9.1 hours (1.2) | overall in lockdown: 9.2h(1.6); strict confinement: 9.3h (1.6); relaxed confinement:9.0h (1.7) | difference before to overall :0.1hour (1.8), p:=0.12; difference strict and relaxed lockdown 0.3, p=0.02 |  |  |  |
| Roy 2020 PP | SC+L | 1065 | 6.86 | 8.18 | 1.32 |  |  |  |
| Watson 2020 (A-D)) | SC+L | 11228 |  |  | sleeping through the night (32% in 2-4 year olds and 50% for 5-7 year olds) = lower than pre-pandemic national data (30% and 60% respectively) |  |  | 33% of parents reporting worse sleep; 7% sleeping better |
| Segre 2020 PP | SC+L | 82 |  |  |  | 61% reported changes in sleep pattern: the majority of them had diculties in falling asleep and woke up many times during the night. They went to bed later than before, but due to school lessons had to wake up at around the same time in the morning. 28% had diculties sleeping and wished to sleep their parents’ bed. |  |  |
| Zhou 2020 | SC+L | 4805 |  | <6h per day:4.5%; 6-8H 59.4%; >8H 36.1% |  |  |  |  |
| Zhou 2020, B. | SC+L | 11,835 |  |  |  |  | Pittsburgh Sleep Quality Index (PSQI): insomnia high scorer (>5): 23.2%. Insomnia associated with depressive symptoms (p<0.001) and anxiety (p<0.001) |  |

SC+L: school closure + lockdown

L: lockdown

#### eTable 12. Health behaviours: 12A. Physical activity (PA), social activity and screentime

| **Study** | **Exposure** | **N** | **Physical activity (hr /week)** | | | **Social activities** | **Screen time (hr/day )** | | | **Social media use (hr/day)** | | |
| --- | --- | --- | --- | --- | --- | --- | --- | --- | --- | --- | --- | --- |
|  |  |  | **pre** | **during lockdown** | **difference** |  | **pre** | **during lockdown** | **difference** | **pre** | **during lockdown** | **difference** |
| Pietrobelli 2020 | SC+L | 41 | May 13 and July 30, 2019. 3.60hr/wk ± 4.25 | March 10, 2020: 1.29 ± 1.44 | −2.30 ± 4.60 :p<0.001 |  | May 13 and July 30, 2019: 2.76 ± 1.64 | March 10, 2020: 7.61 ± 2.13 | +4.85 (SD 2.40) (P<0.001) |  |  |  |
| Dunton 2020 | SC+L | 211 |  | Mean 91.1 (SD = 109.2) min of sitting for school-related activities, 398.5 (SD = 184.6) min of sitting for leisure activity, and 489.4 (SD = 211.5) min of total sitting on the previous day | 36% of parents reported their child had done much less PA in the past 7 days as compared to February 2020;whereas only about 11% of parents reported their child had done much more PA in the past 7 days as compared to February 2020. In contrast, 41% of parents reported their child had done much more sitting in the past 7 days as compared to February 2020, whereas only about 6% of parents reported their child had done much less sitting in the past 7 days as compared to February 2020. |  |  |  |  |  |  |  |
| Lopez Bueno 2020 | SC+L | 860 | mean 198.6 (SD180.9) weekly minutes | strict confinement: 95.5 (123.8); relaxed confinement: 97.8 (121.4) | difference before to lockdown: −102.5 (159.6),p<0.0001 |  | 2.0 (1.6) | strict confinement: 4.9 (2.3); relaxed confinement: 4.8 (2.3) | difference before to during lockdown: +2.9 (2.1); p<0.0001 |  |  |  |
| Roy 2020 PP | SC+L | 1065 |  |  | decreased: 38.6%, remained the same: 37% |  |  | 5.03 |  |  |  |  |
| Widnal 2020 | SC+L | 721-770 |  |  |  |  |  |  |  | High social media use (>=3 hours/day): October 2019: Week-day: girls:42%; boys: 29%; weekend: girls: 62%boys: 46% | April-May 2020: week days: girls: 55%boys: 30%; weekend: girls: 64%boys: 41% |  |
| Watson 2020 (A-D) | SC+L | 11228 |  | worse than before: 47% |  | 47% of parents reported their child did less PA than previously although 24% reported they did more |  |  |  |  |  |  |
| Ellis 2020 | SC+L | 1054 |  |  |  |  | Social media use >3h per day: 31.9%; use >10h per day=2.0% | >3h per day: 77.2%; >10h per day 12.1% |  |  |  |  |
| Zhou 2020 | SC+L | 4805 | PA <30min per day 49.2%; 30-60min 44.3%; >60min 6.5% |  |  |  |  |  |  |  |  |  |
| Gift 2010 | SC | 214 households (269 students) |  |  |  | 69% of students visited other venues during school closure. most students left the home at least once during the closure period to visit routine venues (stores, locations of sports events or practices, restaurants). |  |  |  |  |  |  |
| Johnson 2008 | SC | 220 HH n=355 |  |  |  | At least 1 public location 195 (89%) Grocery stores 97 (44%) Fast food restaurants 77 (35%) Church services 75 (34%) Mall 42 (19%) Parties or sleepovers 33 (15%) |  |  |  |  |  |  |
| Timperio 2009 | SC | 261 |  |  |  | visiting store:113 [43.3%] , visiting family :112 [42.9%], shopping: 101 (38.7%), restaurants: 85 (32.6%) , visiting friends: 79 (30.3%) |  |  |  |  |  |  |
| CDC 2010 | SC | 523 |  |  |  | Overall: 289 (56%) spent time with friends at one another's homes (30%), went grocery shopping (30%), went to fast food restaurants (23%) |  |  |  |  |  |  |
| Effler 2010 | SC | 233 |  |  |  | Overall:172 (74%); most frequent activities: going to sport events; Going to a park or beach; Going to a grocery store; Going to a shopping mall |  |  |  |  |  |  |

SC+L: school closure + lockdown

L: lockdown

SC: school closure (alone)

HH: household

#### 12B. Eating and diet

| **Study ID** | **Exposure** | **N** | **n meals/day** | | | **vegetables intake (servings/day)** | | | **fruit intake(n.serving/day)** | | | **Potato chips (n.serving/day)** | | | **red meat (n.serving/day)** | | | **Sugary drinks (n. /day)** | | | **Eating** |
| --- | --- | --- | --- | --- | --- | --- | --- | --- | --- | --- | --- | --- | --- | --- | --- | --- | --- | --- | --- | --- | --- |
|  |  |  | **pre** | **during** | **diff.** | **pre** | **during** | **diff.** | **pre** | **during** | **diff.** | **pre** | **during** | **diff.** | **pre** | **during** | **diff.** | **pre** | **during** | **diff.** | **during** |
| Pietrobelli 2020 | SC+L | 41 | May 13 and July 30, 2019.: 4.17 ± 0.95 | March 10, 2020: 5.32 ± 1.29 | +1.15 ± 1.56 (p<0.001 | May 13 and July 30, 2019: 1.34 ± 0.74 | March 10, 2020: 1.27 ± 0.69 | −0.07 ± 0.60 (p=NS) | May 13 and July 30, 2019: 1.16 ± 0.74 | March 10, 2020: 1.39 ± 0.70 | 0.23 ± 0.75 (p=NS) | May 13 and July 30, 2019: 0.07 ± 0.24 | March 10, 2020: 0.61 ± 0.83 | +0.54 ± 0.86 (p<0.001) | May 13 and July 30, 2019: 1.80 ± 1.53 | March 10, 2020: 3.46 ± 2.45 | +1.66 ± 2.10 (p<0.001) | May 13 and July 30, 2019: 0.40 ± 0.90 | March 10, 2020: 0.90 ± 1.16 | +0.90 ± 1.16 (P:0.05) |  |
| Lopez Bueno 2020 | SC+L | 860 |  |  |  | 3.2 (2.0) | strict confinement: 3.1 (2.1); relaxed confinement: 2.8 (1.9) | difference before to lockdown: −0.2 (1.6), p=0.0007 |  |  |  |  |  |  |  |  |  |  |  |  |  |
| Roy 2020 PP | SC+L | 1065 |  |  | food intake decreased in 23.6%, increased in 76.4% |  |  |  |  |  |  |  |  |  |  |  |  |  |  |  |  |
| Segre 2020 PP | SC+L | 82 |  |  | 57.3% reported eating more during the lockdown, with an increase in consumption of junk food, snacks, and sweets. 43.9 % reported marked changes in dietary habits particularly consuming more junk food. |  |  |  |  |  |  |  |  |  |  |  |  |  |  |  |  |
| Widnal 2020 | SC+L | students completing both survey ranged from 721-770 |  |  |  |  |  |  |  |  |  |  |  |  |  |  |  |  |  |  |  |
| Watson 2020 (A-D) | SC+L | 11228 |  |  |  |  |  |  |  |  |  |  |  |  |  |  |  |  |  |  | 54% reported eating behaviours unchanged during lockdown, with 32% reported to have worse eating behaviour and 14% reported better eating behaviour |

SC+L: school closure + lockdown

L: lockdown

SC: school closure (alone)

#### eTable 13. Overweight and obesity

| **Study ID** | **Exposure** | **N** | **Body mass index/weight** | | | **Obesity prevalence** | | |
| --- | --- | --- | --- | --- | --- | --- | --- | --- |
|  |  |  | pre | post | difference | pre | post | difference |
| An 2020 | SC+L | 15631 |  | BMI :under Scenarios 1, 2, 3, and 4 with COVID-19, the mean BMIz started at 0.531 (95%CI: 0.515, 0.547) in April 2020, further increased to a value of 0.591 (95%CI: 0.576, 0.607) in September, 0.623 (95%CI: 0.607, 0.638) in August, 0.679 (95%CI: 0.664, 0.695) in October, and 0.718 (95%CI: 0.703, 0.733) in December, and subsequently declined to 0.543 (95%CI: 0.526, 0.560), 0.572 (95%CI: 0.555, 0.588), 0.629 (95%CI: 0.612, 0.645), and 0.685 (95%CI: 0.669, 0.701) in March 2021, respectively. (Fig 2 ) | Relative to the control scenario without COVID-19, Scenarios 1, 2, 3, and 4 were associated with an increase in the mean BMI z-score by 0.056, 0.084, 0.141 and 0.198 respectively | Control scenario:(no school closure): prevalence increased from 13.52% (12.99, 14.06) in April 2020 to 14.77% (14.22, 15.33) in March 2021. | Nnder Scenarios 1, 2, 3, prevalence increased from 13.86% (13.32, 14.41) in April 2020 to 15.41% (14.85, 15.98), 15.74% (15.17, 16.32), 16.45% (5.87, 17.03), and 17.15% (16.55, 17.74) in March 2021, respectively. | Relative to the control scenario without COVID-19, Scenarios 1, 2, 3, and 4 were associated with an increase in childhood obesity prevalence by 0.640, 0.972, 1.676, and 2.373 percentage points respectively. |
| Baysu 2020 | SC+L | 4 |  | weight centile increased from 25-50% centile to 50-75% in all children |  |  |  |  |

SC+L: school closure + lockdown

L: lockdown

SC: school closure (alone)

#### eTable 14. Impacts upon existing conditions

| **Study** | **Exposure** | **N** | **hospital admission** | |  | **Problems in disease management** | **impact on symptoms/severity of the disease** | **Impact on wellbeing/ quality of life** |
| --- | --- | --- | --- | --- | --- | --- | --- | --- |
|  |  |  | **Pre** | **During lockdown** | **Difference** |  |  |  |
| Gelardi 2020 | lockdown-school closure | 120 |  |  |  |  | 85.7-93.3% of parents reported improvements in hearing and auditory symptoms and 44.2-78.2% reported improvements in nasal symptoms |  |
| Martinelli 2020 | lockdown-school closure | 180 | 109/180 (65%) | 29/180 (16.1%) | p<0.01 | For children having immunoregulatory (IM) therapies, 43.1% of parents expressed concerns about the possibility of continuing treatment during COVID-19 pandemic, 7% spontaneously suspended the therapy, and 18.1% postponed their treatments without medical advice. Health-related quality of life scores were similar to previous cohorts |  |  |
| Nastro 2020 | lockdown-school closure | 71 |  |  |  |  | Celiac disease: a non-significant reduced prevalence of all functional gastrointestinal symptoms during lockdown compared to a year previously: from 16 (22.5%) to 12(16.9%): p=0.302. | General Well-being scale mean = 80.13 (64.28–100) indicating an overall good quality of life. 52/71 (73%) did not show anxiety according to the Promis anxiety questionnaires. |
| Christoforidis 2020 | lockdown | 34 |  |  |  | Mean glucose values obtained with sensor did not differ between the two studied periods (168.76 ± 21.87 mg/dl before lockdown versus 170.26± 22.79 during lockdown, p = 0.466). Blood glucose readings were significantly fewer during the lockdown period (7.91 ± 3.45 mg/dl versus 7.41 ± 3.27 mg/dl, p = 0.001) and glucose levels obtained by blood measurements had a significantly higher Coefficient of Variation (CV) in the pre-lockdown period (39.52 ± 5.67% versus 37.40 ± 5.97%, p = 0.011). However, lockdown did not significantly affect glycemic control as mean time in range (TIR) did not differ significantly between the two periods (60.71 ± 13.23% versus 60.50 ± 14.75%, p = 0.872). No significant difference was recorded regarding the total daily dose of insulin required in the two periods (36.24 ± 25.17 U versus 35.80 ± 23.32 U,p = 0.739). No difference was recorded regarding reported carbohydrates during the lockdown period and the preceding 3-weeks period (195.29 ± 106.90 versus 198.06 ±109.13, p = 0.966). Meal schedules changed dramatically during the lockdown period: the percentage of breakfast consumption before 10.00 a.m. fell from 80.67 ± 16.11% before lockdown to 41.46 ± 34.61% (p < 0.001); the percentage of dinner consumption before 10.00 p.m. fell during the lockdown (60.22 ± 26.39% versus53.78 ± 28.97%, p = 0.019) |  |  |
| Darlington 2020 | lockdown | 171 |  |  |  | Any Cancer: For two-thirds (69.6%) of the respondents hospital was no longer considered a safe place. 14 ( 8.1%) reported concern that the response to the COVID-19 situation would lead to suboptimal cancer care or had already led to postponed or cancelled clinic appointments, and several parents were concerned that relapses would bemissed. Parents worried about their own health (81.1%) and about the child contracting the virus from them (89.1%). Parents isolated their chid from immediate family (81.9%). Worries about nurses/careers coming home to vist child could lead to infection( 62.7%) |  |  |
| Marino 2020 | lockdown | 184 parents, 36 young patients |  |  |  | Congenital heart disease: 88% of parents expressed concern regarding the ability of their child’s heart to cope if cardiac symptoms were triggered by the virus .54% of parents felt their child should be isolated from everyone except parents and 70% worried about health care professionals (HCPs) coming into the house.77% worried the hospital was no longer considered a safe place. They worried about their own health (63%) and about the child contracting the virus from them (85%). 84% of planned appointments or surgery were rescheduled. 69% of young patients worried the hospital was no longer a safe place. Some parents ( n=10) were worried surgical procedures being delayed, and the negative consequences for their child. Many parents reported feeling abandoned by the specialist clinical team or the Government (n=34). |  |  |
